# The indirect health impacts of COVID19 restrictions: a strong debate informed by weak evidence

**DOI:** 10.1101/2021.03.16.21253759

**Authors:** Driss Ait Ouakrim, Ameera Katar, Patrick Abraham, Nathan Grills, Tony Blakely

**Author notes:** **Correspondence to:** Dr Driss Ait Ouakrim, Centre for Epidemiology and Biostatistics, School of Population & Global Health, Level 3, 207 Bouverie St., The University of Melbourne, Victoria 3010 Australia. **Contributors and sources** The authors have all been involved in researching aspects of the covid-19 pandemic and response. DA and AK are epidemiologists and have been part of the Victorian Department of Health COVID-19 Public Health Intelligence Team. PA is a health economist involved in evaluating the economic impact of the pandemic. NG is a professor of public health and physician. He has provided expert advice to health agencies on COVID-19 policy, and commentary via science and mass media. TB has been a prominent epidemiological commentator in Australia has been involved in providing modelling underpinning the Victorian Department of Health roadmap out 111 days of lockdown and to achieve elimination. DA, AK and PA designed and run the literature searches in agreement with NG and TB. DA coordinated study selection, data extraction and drafted the paper. All authors debated the findings to reach agreement and contributed to the final version of the manuscript. **Patient involvement** There was no patient involvement in this paper. **Conflicts of Interest** We have read and understood <u>BMJ policy on declaration of interests</u> and have no interests to declare. **Licence** The Corresponding Author has the right to grant on behalf of all authors and does grant on behalf of all authors, an exclusive licence (or non exclusive for government employees) on a worldwide basis to the BMJ Publishing Group Ltd (“BMJ”), and its Licensees to permit this article (if accepted) to be published in The BMJ’s editions and any other BMJ products and to exploit all subsidiary rights, as set out in <u>The BMJ’s licence</u>.

## Abstract

**KEY MESSAGES**

- There has been concern, and much heated debate, on the possible negative effects of restrictions, stay-at-home orders and lockdowns during the COVID-19 pandemic.
- Most published studies on health impacts of restrictions and lockdowns are low quality and often severely biased.
- Focusing on the few studies that approximate a change in restrictions alone (i.e. not the impact of the pandemic per se), we see clear adverse impacts of lockdowns on intimate partner violence and physical activity. Regarding diseases, road traffic crashes decrease, and anxiety increases.
- A discussion driven by science (not politics) is urgently needed on what lockdowns can deliver, their limitations and how to optimally deploy them – along other public health strategies – in the fight against COVID-19.

## Standfirst

### Driss Ait Ouakrim and colleagues

*argue that the vast majority of studies evaluating the unintended effects of COVID-19 restrictions and lockdowns are low quality, prone to severe bias and, therefore, of limited use to health policy making*.

Policies to restrict movements and contact between people have been a common response to control the spread of SARS-CoV-2 in many countries.^1^

The stringency of these measures ranges from recommendations for people to observe physical distancing and self-isolate if symptomatic, to policies imposing strict stay-at-home orders (‘lockdowns’), school closures, business restrictions and border controls.^2^

These restrictive policies, including lockdowns, have been essential to prevent health services being overwhelmed and to save lives.^3, 4^ They have even enabled some countries to eliminate community transmission of the virus.^5^ What remains unclear, however, are the potential unintended health harms associated with such policies.

Public health authorities and commentators have raised the alarm – as early as the first lockdowns in Wuhan – on the indirect health harms that might result from lockdown policies.^6^ The potential mental health impacts, ranging from loneliness and anxiety through to severe depression and even suicide, have been raised as major concerns.^7, 8^ Other proposed harmful effects have included cardiac conditions, delayed screening, presentation or treatment for cancers, intimate partner violence, increased consumption of ultra-processed foods, alcohol, tobacco and illicit substances.

But what is the actual quantitative evidence on these harms? To answer this, we argue, requires thinking counterfactually before considering approaching the plethora of published studies on this topic.^9^

## Counterfactual reasoning and the net health effect of lockdowns

Health impacts during lockdown are a blend of the effect of the lockdown itself (the relevant target for causal inference) and the general effect of the pandemic. For example, people’s mental health may be adversely impacted by fear and uncertainty of the pandemic, plus any additional impact of restrictions per se. We want to identify the unintended causal effects (good and bad) of restrictions all other things held constant. Whilst no randomised trials have approximated this, we use natural experiments^10^, namely:

- Comparisons over time between residents in the same place during the pandemic, with varying restrictions (i.e. changing exposure) but otherwise similar circumstances (i.e. an absence of time varying confounding, either by design or statistical adjustment).
- Comparisons across places (e.g. states of a country) of similar populations with similar SARS-CoV-2 infection rates and similar other circumstances, but variation in levels of restriction.

That is, we need to find populations that are exchangeable with each other, and only vary in terms of the level of restrictions. This rules out many cross-sectional surveys. Furthermore, many studies that purport to show impacts of restrictions contain significant biases. For example, people cannot be blinded to their own lockdown status which means that subjective measures such as feelings of anxiety are prone to recall bias whilst rapidly conducted internet recruitment surveys are typically not representative of the population (i.e. selection bias).

## State of the evidence

To hold such a counterfactual ruler up against the available evidence, we searched the literature for studies meeting or approximating the counterfactual described above. First, we selected risk factors and diseases that have a plausible independent association with COVID-19 policy responses, and that are likely to contribute to a non-negligible health burden: five risk factors (alcohol consumption, body mass index, intimate partner violence, physical activity and tobacco smoking); five health seeking behaviours (screening for melanoma, breast, cervical, colorectal and lung cancer); and ten disease/injury groups (anxiety and depression, chronic obstructive pulmonary disease (COPD), falls, ischemic heart disease (IHD), self-harm, stroke, suicide and road transport injuries).

The vast majority of the existing literature up to 21 January 2021 was low quality and/or contained strong bias (see Appendix for details), leaving only 42 articles. Of these, 28 were comparisons of before with during the pandemic – but were still too weak due to having two or more potentially substantial biases (confounding, measurement error, or selection). This left only 14 studies meeting our strict criteria, and still the majority of these were before-COVID-19 to during COVID-19 comparisons prone to residual confounding.

## Consequences

Starting with the fourteen ‘best studies’, wherever possible we converted the study finding into a percentage change for the equivalent of severe restrictions (typically referred to as stage 3 or 4 lockdown) compared to minor restrictions only (see Appendix for details). Where the conversion was not possible, the percentage change was plotted as reported in the study. Figures 1 shows these results for risk factors and diseases.

**Figure 1.**
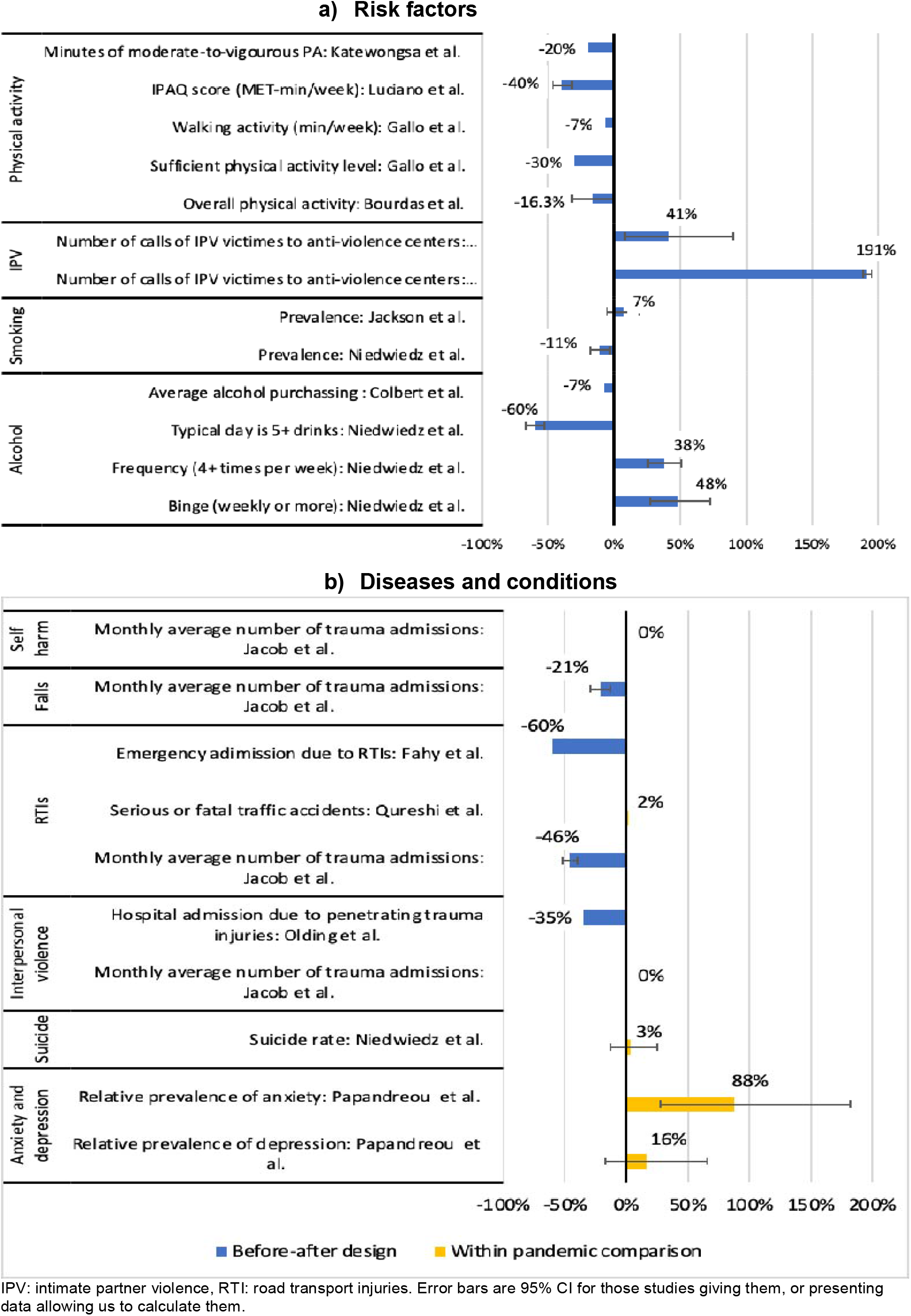
Lockdown effect estimates from studies using natural experiment or before-after designs.

### 1) Higher risk of intimate partner violence

Two well designed before-after studies conducted in Spain^11^ and Italy^12^ showed a sharp increase in the number of women reaching out for help to anti-violence centres. The analyses demonstrate a surge in the number of calls (ranging from 41% to 191%) to these centres from women experiencing violence or in a threatening environment during lockdown periods. Both studies were able to access data from all centres operating in their respective countries, which reduces the level of potential measurement error and virtually eliminates risk of selection bias. The findings are in line with many reports and initiatives highlighting the higher risk of violence against women during the COVID-19 crisis, and the need to prevent what has been termed as the “shadow pandemic”.^13^

### 2) Lower physical activity

As one would expect, restrictions and lockdowns lead to a substantial drop in the average level of physical activity.The four studies^14-17^ we include used different measures of physical activity but all reported a reduction (ranging from 7% to 40%). The analyses were based on specific population groups (e.g. medical student, or patients with chronic condition) which might limit the generalisability of their results. What we don’t know, however, is how lower physical activity during lockdowns will affect future population health outcomes. Modelling studies are needed in this area to estimate the potential effects of reduced physical activity and help design and evaluate future mitigation strategies.

### 3) No convincing impact on alcohol and tobacco use

The studies reporting on the effect alcohol consumption and tobacco smoking during lockdown periods provide conflicting results for these two risk factors. Smoking habits do not seem to have been impacted by the lockdowns. Jackson et al.^18^ reported a 7% increase in smoking prevalence (95% CI: 5%-19%), while Niedwiedz et al.^19^ showed a reduction of 11% (95% CI: −18% to −3%). There was similar uncertainty around alcohol consumption. Colbert et al.^20^ identifies a 7% reduction in the alcohol purchasing during the lockdown, while Niedwiedz et al.^19^ found changing patterns in consumption that might cancel out any effect.

### 4) Reduced number of injuries due to falls and road traffic

A clear positive health effect of restrictions and lockdown policies seems to be the substantial reduction in the number of road traffic injuries as shown by two studies analysing the number of emergency department admissions in Ireland and Australia.This effect likely results from the reduction in mobility induced by lockdowns and minimal economic activity.^21^ However, the US study by Quershi et al.^22^ which analysed data from the Statewide Traffic Accidents Record in Missouri showed no effect of the lockdown on the number of road traffic accidents resulting in serious or fatal injuries. Which suggest that the wider context in which a lockdown is implemented might determine how it impacts road traffic injuries, e.g. a highly car dependent culture may actually use road travel to socially isolate.

Other beneficial effects include a reduction in trauma admission due to falls. Jacob et al.^23^ reported a 21% reduction observed in a large trauma centre in Australia compared to pre-lockdown levels. This also seems a very plausible consequence of lockdowns, if indeed people are mandated to stay at home and limit their movement to the strict minimum.

### 5) Likely higher level of anxiety

We identified only one study investigating the effect of lockdown on anxiety that met our counterfactual criteria. Papandreou et al.^24^ compared the levels of anxiety using two parallel surveys: in Spain, while the country was experiencing a strict lockdown, and in Greece where minimal restrictions were in place. The prevalence of anxiety in the Spanish population was 88% (95% CI: 27%-182%) greater than in the Greek population. The potential risk of confounding from the pandemic itself is likely to be low, under the assumption that the pandemic induces a similar level of anxiety in the two populations. However, the study does not adjust for any pre-pandemic variation in anxiety levels.

### 6) Little evidence of effect on depression, self-harm and suicide

The three studies reporting on depression^24^, self-harm^23^ and suicide^19^ suggest that lockdowns have no immediate effect on the prevalence of these conditions. While this finding seems plausible in the short term and for a given period of lockdown, it does not provide definitive evidence of absence of effect. First, there remains uncertainty on how the experience of lockdown might translate in the long-term – in particular on prevalence of depression; and second, how repeated periods of restrictions and lockdowns – as has been experienced in many countries – might impact these three conditions and mental health in general.

#### Box 1

**Disruptions in cancer care and prevention**

- There is strong and consistent evidence, from a variety of countries, that disruptions related to the pandemic have led to substantial reductions in cancer screening, diagnoses and treatment. Denmark, for example, saw a 33% (95% CI:26%-40%) reduction in incident cancers between March and May 2020 compared to the same months in the previous five years.^25^ In the Netherlands, cancer diagnoses (across all sites, excluding skin cancer) were 20% lower than in 2019.^26^ In the United States, the weekly number of newly identified cancers declined by 46% for 6 cancer types combined (i.e. breast, colorectal, lung, pancreatic, gastric, and oesophageal).^27^ Similar declining trends in referrals and diagnosis have been consistently reported in other countries and for individual cancers.^28-30^ However, we are not aware of studies that separate out the effect of restrictions per se on cancer service utilisation, from that due to the wider pandemic.
- Modelling studies have also evaluated the potential impact of pandemic-related disruptions to cancer treatment and prevention. In the UK for example, a 3 month delay in referral/treatment is expected to result in 4,500 lives lost over a period of 10 years (Sud et al.).
- These delayed presentations result from both increased pressure that the pandemic has placed on health services and the fear of attending them. How much of any contemporaneous impact is due to the causal impact of restrictions, as opposed to fear of contacting people with SARS-CoV-2 and health services capacity being exceeded, is unknown. Moreover, there is a case that restrictions and lockdowns – by reducing future case loads – will in the medium term increase screening compared to a counterfactual of ongoing high case loads disrupting health services.
- In most countries, healthcare resources such as staff were reassigned to COVID-19 leading to negative effects on cancer care and other chronic diseases.^31^ However, even where countries didn’t suspend their national cancer screening programmes, uptake decreased possibly out of fear of catching Covid19 or due to assumptions about the lack of capacity in the health system, or presumed closure.
- The specific role that restrictions played in these disruptions to healthcare is less clear. The few studies that investigated the specific contemporaneous effect of the lockdown were likely confounded by the broader effect of the pandemic.

## What is beyond the counterfactual framework?

Relaxing our criteria to include weaker study designs (before-after studies with two or more major likely biases), we considered 32 additional studies. Summaries of their main features and findings are provided in the online appendix. These consistently show that the risk factors worsen during (but perhaps not because of) lockdowns.

The same negative effects were shown for many diseases and conditions, apart from road traffic injuries, for which a substantial reduction was reported, consistent with the results from the higher quality studies.

For six out of 10 conditions considered, the primary outcomes were access to health services, which represents a further limitation to these studies. Health seeking behaviour might not accurately represent change in incidence. For example, mild symptoms of ischaemic heart disease or COPD might be ignored as a result of the pandemic out of fear of going to hospital.

## Is the cure, indeed, worse than the disease?

The positive effect of restrictions containing a deadly pandemic is clear. But there is no doubt measures such as lockdowns are highly disruptive at the time and can have profound social, health and economic consequences. They also force policy makers and society as a whole to address challenging ethical and philosophical questions.^32^ It is well established that public health interventions in general can have unintended harmful effects, reinforce health inequities or even worsen the negative outcomes they set out to address.^33^ Our data suggests lockdown’s can have both negative and positive consequences such as decreasing road traffic injuries, yet increasing risk of intimate partner violence and reducing physical activity.

However, in the case of the COVID-19 pandemic, what should have been a rigorous and evidence-based discussion between public health experts and policy makers^34^ about how to best implement a preventive strategy to protect population health, turned into a sterile and highly politicised debate^35^ between those who support the lockdowns – seeing them as the only way to tackle the pandemic in the absence of effective treatment or vaccine – and those claiming that their unintended consequences are so severe that they cause more harm than the pandemic itself.^36^

Accordingly, some governments imposed long and unnecessarily harsh lockdowns, even where the harm seemingly outweighed the benefit (e.g. India, Philippines). Other governments perhaps overestimated the negative impact of lockdowns and only used them as a last resort (e.g. France, Germany) or ruled them out as a preventive strategy (e.g. Sweden, Brazil).

Using a counterfactual filter on studies we found little quality evidence to suggest the cure is worse than the disease. However, absence of quality evidence isn’t necessarily evidence of absence of negative (or positive) effect. Clearly, we need more quality, natural-experiment data, along with data on delayed health effects, including those resulting from social determinants such education and employment. From our review and analysis, we believe much of this information exist – hidden in governmental and private databases – but has not (yet) been analysed or published.

We urge researchers to try and disentangle the causal impact of lockdowns and restrictions for ‘background’ COVID-19, to hopefully allow a more scientific (and less political) discussion about what they can deliver, their limitations and how to optimally deploy them – along other public health strategies – in the fight against COVID-19.

## Data Availability

N/A

## References

1. Han E, Tan MMJ, Turk E, Sridhar D, Leung GM, Shibuya K, et al. Lessons learnt from easing COVID-19 restrictions: an analysis of countries and regions in Asia Pacific and Europe. The Lancet. 2020;396(10261):1525–34.

2. Cheng C, Barceló J, Hartnett AS, Kubinec R, Messerschmidt L. COVID-19 Government Response Event Dataset (CoronaNet v.1.0). Nature Human Behaviour. 2020;4(7):756–68.

3. Chaudhry R, Dranitsaris G, Mubashir T, Bartoszko J, Riazi S. A country level analysis measuring the impact of government actions, country preparedness and socioeconomic factors on COVID-19 mortality and related health outcomes. EClinicalMedicine. 2020;25:100464.

4. Hsiang S, Allen D, Annan-Phan S, Bell K, Bolliger I, Chong T, et al. The effect of large-scale anti-contagion policies on the COVID-19 pandemic. Nature. 2020;584(7820):262–7.

5. Baker MG, Wilson N, Blakely T. Elimination could be the optimal response strategy for covid-19 and other emerging pandemic diseases. BMJ. 2020.

6. J B, S G, M K. The Great Barrington Declaration 2020 [Available from: https://gbdeclaration.org/.

7. Brooks SK, Webster RK, Smith LE, Woodland L, Wessely S, Greenberg N, et al. The psychological impact of quarantine and how to reduce it: rapid review of the evidence. The Lancet %@ 0140-6736. 2020.

8. Courtet P, Olié E, Debien C, Vaiva G. Keep socially (but not physically) connected and carry on: preventing suicide in the age of COVID-19. The Journal of clinical psychiatry. 2020;81(3):0-%@ 0160–6689.

9. Mansfield KE, Mathur R, Tazare J, Henderson AD, Mulick AR, Carreira H, et al. Indirect acute effects of the COVID-19 pandemic on physical and mental health in the UK: a population-based study. The Lancet Digital Health. 2021.

10. Craig P, Katikireddi SV, Leyland A, Popham F. Natural Experiments: An Overview of Methods, Approaches, and Contributions to Public Health Intervention Research. Annual review of public health. 2017;38:39–56.

11. Beigelman M, Vall Castelló J. COVID-19 and help-seeking behavior for intimate partner violence victims. IEB Working Paper 2020/13. 2020.

12. Lundin R, Armocida B, Sdao P, Pisanu S, Mariani I, Veltri A, et al. Gender-based violence during the COVID-19 pandemic response in Italy. Journal of global health. 2020;10(2).

13. UN Women. The Shadow Pandemic: Violence against women during COVID-19 2020 [Available from: https://www.unwomen.org/en/news/in-focus/in-focus-gender-equality-in-covid-19-response/violence-against-women-during-covid-19.

14. Bourdas DI, Zacharakis ED. Impact of COVID-19 Lockdown on Physical Activity in a Sample of Greek Adults. Sports (Basel). 2020;8(10).

15. Gallo LA, Gallo TF, Young SL, Moritz KM, Akison LK. The Impact of Isolation Measures Due to COVID-19 on Energy Intake and Physical Activity Levels in Australian University Students. Nutrients. 2020;12(6).

16. Katewongsa P, Widyastaria DA, Saonuam P, Haematulin N, Wongsingha N. The effects of COVID-19 pandemic on physical activity of the Thai population: Evidence from Thailand’s Surveillance on Physical Activity 2020. J Sport Health Sci. 2020.

17. Luciano F, Cenacchi V, Vegro V, Pavei G. COVID-19 lockdown: physical activity, sedentary behaviour and sleep in Italian medicine students. Eur J Sport Sci. 2020:1–22.

18. Jackson SE, Garnett C, Shahab L, Oldham M, Brown J. Association of the Covid-19 lockdown with smoking, drinking, and attempts to quit in England: an analysis of 2019-2020 data. medRxiv. 2020.

19. Niedzwiedz CL, Green MJ, Benzeval M, Campbell D, Craig P, Demou E, et al. Mental health and health behaviours before and during the initial phase of the COVID-19 lockdown: longitudinal analyses of the UK Household Longitudinal Study. Journal of Epidemiology and Community Health. 2020:jech-2020–215060.

20. Colbert S, Wilkinson C, Thornton L, Richmond R. COVID-19 and alcohol in Australia: Industry changes and public health impacts. Drug Alcohol Rev. 2020;39(5):435–40.

21. De Vos J. The effect of COVID-19 and subsequent social distancing on travel behavior. Transportation Research Interdisciplinary Perspectives. 2020;5:100121 %@ 2590-1982.

22. Qureshi AI, Huang W, Khan S, Lobanova I, Siddiq F, Gomez CR, et al. Mandated societal lockdown and road traffic accidents. Accident Analysis & Prevention. 2020;146:105747.

23. Jacob S, Mwagiru D, Thakur I, Moghadam A, Oh T, Hsu J. Impact of societal restrictions and lockdown on trauma admissions during the COVID-19 pandemic: a single-centre cross-sectional observational study. ANZ journal of surgery. 2020.

24. Papandreou C, Arija V, Aretouli E, Tsilidis KK, Bulló M. Comparing eating behaviours, and symptoms of depression and anxiety between Spain and Greece during the COVID-19 outbreak: Cross-sectional analysis of two different confinement strategies. European Eating Disorders Review. 2020;28(6):836–46.

25. Skovlund CW, Friis S, Dehlendorff C, Nilbert MC, Mørch LS. Hidden morbidities: drop in cancer diagnoses during the COVID-19 pandemic in Denmark. Acta oncologica. 2021;60(1):20–3.

26. Dinmohamed AG, Visser O, Verhoeven RHA, Louwman MWJ, van Nederveen FH, Willems SM, et al. Fewer cancer diagnoses during the COVID-19 epidemic in the Netherlands. Lancet Oncol. 2020;21(6):750–1.

27. Kaufman HW, Chen Z, Niles J, Fesko Y. Changes in the Number of US Patients With Newly Identified Cancer Before and During the Coronavirus Disease 2019 (COVID-19) Pandemic. JAMA Network Open. 2020;3(8):e2017267.%@ 2574-3805 %[3/9/2021 %U https://doi.org/10.1001/jamanetworkopen.2020.17267.

28. D’Ovidio V, Lucidi C, Bruno G, Lisi D, Miglioresi L, Bazuro ME. Impact of COVID-19 Pandemic on Colorectal Cancer Screening Program. Clinical colorectal cancer. 2020.

29. Morris EJ, Goldacre R, Spata E, Mafham M, Finan PJ, Shelton J, et al. Impact of the COVID-19 pandemic on the detection and management of colorectal cancer in England: a population-based study. The Lancet Gastroenterology & Hepatology. 2021.

30. Park JY, Lee YJ, Kim T, Lee CY, Kim HI, Kim JH, et al. Collateral effects of the coronavirus disease 2019 pandemic on lung cancer diagnosis in Korea. BMC Cancer. 2020;20(1):1040.

31. WHO Newsroom. COVID-19 significantly impacts health services for noncommunicable diseases [Available from: https://www.who.int/news/item/01-06-2020-covid-19-significantly-impacts-health-services-for-noncommunicable-diseases.

32. Chu IY-H, Alam P, Larson HJ, Lin L. Social consequences of mass quarantine during epidemics: a systematic review with implications for the COVID-19 response. Journal of travel medicine. 2020;27(7):taaa192.

33. Bonell C, Jamal F, Melendez-Torres GJ, Cummins S. ‘Dark logic’: theorising the harmful consequences of public health interventions. J Epidemiol Community Health. 2015;69(1):95–8.

34. Lorenc T, Oliver K. Adverse effects of public health interventions: a conceptual framework. J Epidemiol Community Health. 2014;68(3):288–90.

35. Goodman SW, Pepinsky TB. Partisanship, health behavior, and policy attitudes in the early stages of the COVID-19 pandemic. SSRN Electron J. 2020.

36. Melnick ER, Ioannidis JPA. Should governments continue lockdown to slow the spread of covid-19? BMJ. 2020;369:m1924.

