## Supplementary Files for "The indirect health impacts of COVID19 restrictions: a strong debate informed by weak evidence"

### Introduction

In this supplement we expand on three areas:

1) The literature search methodology and study selection criteria
2) Narrative and tabular description of the included studies for
   a. Risk factors
   b. Conditions and diseases
3) Details of the literature search strategies, search terms and PRISMA diagrams

### 1. Literature search methodology and study selection criteria

We conducted a scoping review^1^ of the studies published from 1 January 2020 to 21 January 2021.

PubMed and Google Scholar were searched for peer-reviewed articles, preprints and institutional reports, using the risk factors and conditions listed in the main manuscript in combination with different expressions describing Covid19 control policies.

We included studies conducted in OECD countries and China. The highest quality studies (what we term Category 1a) were either a times series study that included variation in restrictions or lockdowns during the pandemic, or a comparison across places with varying restrictions or lockdowns. Category 1b studies were before-after analyses in one region or jurisdiction, where the ‘before’ measure preceded the pandemic meaning they are unable to disentangle the effects of the pandemic per se versus social restrictions and lockdowns. These category 1b studies comprised the majority of Category 1 studies. Category 2 studies were other studies meeting our initial inclusion criteria, but either not meeting the Category 1 criteria or containing two or more serious biases (from two or more of confounding ^[1]^, measurement error or misclassification ^[2]^, and selection bias^[3]^). Team consensus was used to determine bias.

Single studies often contributed more than one measure of interest (e.g. smoking and alcohol). Each measure of interest was considered separately for bias, with many measures of interest being rejected from category 1 (e.g. quit attempts were ascertained for the last year, but the lock down was only the last month – a severe example of measurement error). The main text of this paper focuses on possible bias in those study’s that made Category 1.

We extracted and summarized key information from the included studies, including country, design and published effect size. We attempted to classify studies into four possible stages of restriction, as described in Table S.1 However, most studies just compared lockdown to none, where lockdown was usually equivalent to stage 3 in our classification of states. Ideally, studies that reported data from across the stages could help discriminate linear dose-response relationships from stage 0 to 4, or a step function for stages 0 to 2, compared to stages 3 and 4 (given stay at home orders occur from stage 2 to 3); however very few such studies were found.

For those studies we included in Category 1, we converted (where possible) their study results into a common metric – the percentage (i.e. relative, not percentage point) change for the equivalent of stage 3 or 4 versus stage 0. For outcomes measured using a continuous scale (e.g. measures of depressive symptomatology) we first estimated what percentage of the study population might be classified with a condition (e.g. using standard cut-points on a depression inventory for moderate to severe depression) and then estimated how the difference reported in the study (e.g. change in mean score) would change the proportion of the study classified as ‘diseased’.

We did not explicitly search for on-line data series where it was not reported in a publication captured by our search criteria. For example, we did not search Government published data on road traffic crash rates, plot it and analyse it by level of restriction over time, and report on these analyses ourselves. However, we strongly encourage further studies of this type.

Table S.1: Characteristics of restriction stages

|  | **Stage 4** | **Stage 3** | **Stage 2** | **Stage 1** |
| --- | --- | --- | --- | --- |
| **Oxford Stringency Index †** | 70 to 100 | 50-70 | 31-50 | 0-30 |
| **International / State Borders** | Completely closed | Closed, repatriation flights only | Open to citizens | Open |
| **Workplaces** | Personal services closed, strict restrictions on transport, logistics and manufacturing. Construction stopped | Work from home if you can, restrictions apply for personal services, construction, transport, logistics and manufacturing. | Work from home if you can, restrictions apply for personal services, transport, logistics and manufacturing | Work from home if you wish |
| **Visitors to Home** | No visitors at all, except for compassionate reasons | 5 people can visit one household | Max 10 guests, good physical distancing, | Nil restrictions |
| **Time away at home** | 1 hr max daily for exercise, shopping, or care. Masks to be worn at all time outside of home | Only for essential work, exercise, shopping, caring. Masks essential outside of the home | No restrictions | No restrictions |
| **Retail** | Only essential retail open, i.e., supermarkets, pharmacies. Coffee shops/ restaurants open for take away only | Masks at shops, minimal retail open, no more than one person per 4sq metres of floor space | Masks at shops, no more than one person per 4sq metres of floor space | All open, no more than one person per 4sq metres of floor space |
| **Public Transport** | Only for essential workers, must wear masks | Only for essential workers, must wear masks | Reducing capacities | All open |
| **Educational Institutions** | All closed | Schools only open for children of essential workers, universities closed. | Schools open, universities closed. | All open |
| **Gathering Limits** | 10 people max for funerals and weddings, strict physical distancing + masks: no more than one person per 4sq metres of floor space | 50 people indoors with physical distancing +masks, 100 outdoors with physical distancing, no more than one person per 4sq metres of floor space | 100 people indoors with physical distancing, 200 outdoors with physical distancing +masks | 250 people indoors with physical distancing, 500 outdoors with physical distancing |

† Cut-points determined by comparing NZ and Australian data with known ‘stages’ in these countries. However, it must be noted that the Oxford Stringency Index is that or the most restricted area in each country at that time; a lock down in just one region of the country means the whole country is classified with a high stringency score.

Table S.2: Risk of bias of high quality studies included in main analysis

| Study design | Within pandemic time series or comparison over place study | | Before compared to during pandemic study | |
| --- | --- | --- | --- | --- |
| Bias potential | None or one potential major source of bias  [graphed in Figure 1] | Two or more potential major source of bias from two or more types (confounding, measurement, selection bias) | None or one potential major source of bias  [graphed in Figure 1] | Two or more potential major source of bias from two or more types (confounding, measurement, selection bias) |
| Number of studies | 3 | 0 | 7 | 32 |

### 2. Narrative and tabular summary of the included studies

#### Risk factors

##### Alcohol

Using the UK Household Longitudinal Study Niedwiedz et al (2020)^2^ determined differences in alcohol consumption in three historic waves centred on 2016, 2017 and 2018 respectively, with data collected April 2020 during a lockdown. They use a multilevel model with survey year included to capture time trends, and estimate a relative percentage change in three alcohol-related metrics as shown in Figure 1 in the article. However, they did not report changes in average consumption.

Colbert et al (2020) ^3^ report early data for alcohol purchasing that can be tracked using bank eftpos data. There is one key weakness – possible misclassification of actual alcohol consumption if people are either stockpiling or displacing purchasing from cash to electronic means of purchase. But selection bias and confounding were unlikely, so we included this study as Category 1b. The authors report a 34% increase in alcohol purchasing in week 1 (ending 27 March 2020) and 28% purchasing in week 2 (ending 3 April; however this was an error as the source data was -10% [28% was for bottle shops, which together with alcohol services with a 71% reduction gives -10% overall]) and a decrease of 13% in week 3 (ending 10 April). Using source the correct source data to replicate the authors analysis for the six week period from 21 March to 1 May, we found that there was in fact an average 6.2% reduction in alcohol purchasing - which adjusting for annual demographic growth of 1.3% (<https://www.abs.gov.au/statistics/people/population/national-state-and-territory-population/latest-release>) is a 7.4% decrease.

Tran et al (2020) ^4^ undertook an anonymous online survey, asking people if they had increased or decreased alcohol consumption. No estimate of amount or pattern of drinking was elicited, meaning the study cannot be used in quantification. It is also prone to selection bias given the sampling frame, and recall bias as separate pre-lockdown data was not collected (i.e. given the non-blinded lockdown exposure, differential misclassification of alcohol consumption in past (pre-lockdown) compared to during lockdown is likely). Nevertheless, participants were twice as likely (20.9%) to say they were drinking more than they used to compared saying less than the used to (10.5%) – but the largest category was no change (43.9%).

Similar to Tran et al, Rolland et al (2020)^5^ conducted a web-based survey to elicit recalled changes in alcohol consumption – which found a 24.8% relative reduction in alcohol consumption during lockdown in France. This study has the same two serious limitations as above, removing it from Category 1 to Category 2, namely: drinking rates and patterns were not quantified, as well as potential differential recall of pre-lockdown alcohol consumption.

Dumas et al (2020)^6^ conducted an online survey for Canadian youth, aged 16-18, and found that the percentage of the surveyed adolescents who self-reported alcohol use “in the past 3 weeks before the COVID-19 crisis” and “since the COVID-19 crisis (e.g., the past 3 weeks)” did not change significantly from pre- COVID to post-COVID (28.6% to 30.1%, p = 0.23; no CI reported). Additionally, binge drinking rates decreased from 15.7% to 9.8% (difference of 5.9%). Pre Covid-19, 15.7% of respondents engaged in binge drinking, compared with 9.8% during lockdown. However, this study suffers from two serious potential biases: a) selection bias, as it was an online Instagram recruitment (not population representative), and b) measurement error in that self-report was used, and for further back in time for the pre-COVID crisis. For these reasons, we relegated this study to Category 2.

*Average daily alcohol intake: Whilst Niedwiedz et al^2^ found evidence of changing patterns in consumption, these patterns of consumption change may cancel out to nil change in daily average intake. The other category 1 level evidence, Corbet et al, upon correction of reported data and reanalysis, found a modest decrease. Our conclusion is therefore nil or little change, but with moderate uncertainty. For example, it would be reasonable to assume a best estimate of 0% change in average alcohol intake, but with +/- 10% SD … pending further evidence that either or both shifts the best estimate and narrows uncertainty. Further, it is unknown that if alcohol intake does change during lockdowns, whether any change persists after the end of lockdown.*

Body weight and BMI

Pellegrini et al (2020)^7^ undertook a cohort study of 150 overweight and obese people in an outpatient weight loss programme. Participants were instructed to weight themselves daily, and using these reported weights there was an average 1.5kg weight gain and 0.58 kg/m^2^ BMI increase from pre-lockdown to 1-month post-commencement of lockdown. The fact that weight increased, when the intent was weight-loss, does speak to the obesogenic influence of lockdown among people already with excess weight enrolling in an outpatient programme. However, there is no counterfactual comparison – what would have the weight change been in these people in the programme with no lockdown? Second, the results only speak to the sub-population who are overweight and obese enrolling in outpatient weight loss programs. It is therefore category 2 contextual information – supporting the hypothesis that lockdowns can lead to weight increase.

He et al^8^ report on self-reported changes in body weight among 339 respondents to social media recruitment, where each respondent was asked to self-report their weight five weeks prior to lockdown (27 Jan), and in the month of lock down. As a non-representative survey with potential selection bias towards those people who had put on weight (i.e. due to interest in responding), and using self-report, the results are not high quality. Nevertheless, individuals with a BMI less than 25 pre-lockdown had 2.2kg and 1.7kg increases in weight for females and males respectively (no CI given, but P<0.01 for both before after comparisons). For people with a BMI greater than 25, there was a 0.9 kg increase for females (p=0.042) and a 0.09 decrease for males (p=0.003). Whilst not a rigorous study design, the results suggest weight increased for initially normal weight people.

*Body weight summary: There is no reliable estimate of impact on body weight. Nor would one be easy to detect for lockdowns of usually short duration, due to slow changes in body weight – theoretically making any impact from lock-downs small and unlikely to be detectable.*

*Theoretically, again, even if there is no (detectable) impact of lockdowns on body weight, this does not rule out an effect on body weight of lesser restrictions (of likely longer duration). For example, for a locality not in lockdown, but where people are encouraged to work from home as much as possible, body weight may change among these people if dietary intake and/or physical activity changes as a result.*

##### Intimate partner violence (IPV)

Two studies investigating the effect of restriction on the level of intimate partner violence. Only two studies met the inclusion criteria. Both studies reported data on the number of calls to women support services related to intimate partner violence during lockdown periods.

Beigelman et al (2020)^9^ analysed rates of emergency calls associated with IPV during each month of the lockdown in each province in Spain, compared with 2019 data. The analysis showed that the lockdown was associated with a 40.7% increase in emergency calls from IPV victims. The difference in difference co-efficient showed an increase of 3.51 calls per 100,000 (standard error 0.37) during the lockdown period (which converted to the percentage scale gives a 95% CI about 40.7% of 32.3% to 49.1%).

Lundin et al (2020)^10^ compared number of calls from women to 58 anti-violence centres across Italy during two periods of lockdown (between 2 March and t April 2020 and between 6 April and 3 May) to the monthly average number of calls received by the same centres between 2016 and 2018. The monthly average of calls was stable between 2016 and 2018 (mean value 1306 women, 95% CI 1255 to 1357). The analysis showed a 191% increase (derived 95% CI: 188%-195%) in the number of calls compared 2016 and 2018. Women with previous history of calls represented less than a third of callers in 2016-2018 but over two third in 2020. Whilst callers were of course not blinded to the lockdown status, there was no other likely major bias.

*Intimate partner violence summary. Based on two studies during lock downs in Spain and France user emergency calls regarding IPV, an increase seems highly likely. The best estimate might be reasonably the average. Given the percentage values, and that they are large and variable, a log transformation is appropriate to estimate a geometric mean (ratio 2.02, or a 102% increase, or an approximate doubling). The uncertainty, especially considering likely generalizability to other settings, is likely large. It would seem sensible to set the SD as half the difference (log scale) in the two input studies, putting a 95% uncertainty interval at a ratio of 0.99 to 4.12 ( or a two-fold increase with uncertainty ranging from nil to four-fold increase).*

##### Physical activity

Six studies met our inclusion criteria:

Gallo et al.^11^ evaluated the impact of isolation (eg. physical and social distancing, work or study from home where possible) measures on diet and physical activity pattern in Australian undergraduate biomedical students approximately one week after transition to online learning. Students aged under 27 years were asked to complete The Automated Self-Administered Dietary Assessment Tool^12^ and to recall all foods, drinks, and supplements consumed in the previous 24 hours. Participants also completed the Active Australia Survey^13^ to estimated leisure-time physical activity over the preceding week. The study was initiated in 2018, the authors were therefore able to compare 2020 survey results to the findings from the two previous years. The survey showed that energy intake increased by 20% for females students in the 2020 class, but not for males. The level of physical activity decreased by 30% for both sexes during the pandemic.

The study was classified in Category 2 as no counterfactual comparison was conducted. The findings apply to a non-representative sample and are exposed to confounders as well as a high-risk of selection and recall bias.

Tison et al.^14^ conducted a before-after study using smartphone data collected between 19 January and 1 June 2020 from over 455k users of a popular health and wellness app to analyse daily step counts in a selection of countries. They calculated mean number of daily steps between 19 January and 11 March 2020 (i.e. pre-pandemic) and between the 12 March and first of June (during pandemic). They determined the percentage change in mean number of steps for the included jurisdictions by comparing the two time periods. Worldwide, within 10 days of the pandemic declaration, there was a 5.5% decrease in mean steps, and within 30 days, there was a 27.3% decrease in mean steps. The results suggest a reduction in physical activity but show wide regional variations in the physical activity measure used and in the timing and rate of that change reflecting differences in national policy responses to COVID-19. However, this study was classified as category 2 as there was no attempt to analyse the data by whether people were in lock down or some varying magnitude of imposed restrictions (versus ‘just’ exposure to the pandemic whish may reduce PA as people seek to have less social contact regardless of imposed lockdowns). There was also a risk of selection bias if people using the app responded to differently to lockdowns and restrictions than all people.

López-Sánchez et al. conducted a cross-sectional survey to evaluate the levels of physical activity in adults with chronic conditions before and during the lockdown put in place in Spain in March 2020. The authors found a significant decrease in moderate-intensity physical activity during the lockdown (mean difference: 30 min/day). Vigorous-intensity physical activity saw decreases of 15.5 min/day for males and 2.6 min/day for females. The analysis was based on a convenience sample of participants recruited through social media between 1 April 2020 and 1 May 2020 (i.e. after the lockdown was put in place). There is there a high risk of measurement error, in addition of selection bias as the study was conducted among people with chronic conditions. Furthermore more, no counterfactual comparison was conducted. The study was therefore classified as category 2.

Luciano et al.^15^ surveyed 6^th^ years medical students to evaluate their level of physical activity, sedentary behaviour and sleeping habits before (October- November 2019) and during the strict lockdown that was implemented in Italy between 9^th^ of March and 3^rd^ of April 2020. Physical activity was evaluated using the International Physical Activity Questionnaire Short Form (IPAQ-SF) and IPAQ score expressed as metabolic equivalent minutes per week (MET-min/week). Sleep and rest were evaluated by means of selected questions from Pittsburgh Sleeping Quality Index Questionnaire (PSQI). The authors found a during lockdown, with a reduction of 628 MET-min/week, which represent a 40% reduction compared to pre-lockdown estimate.

Bourdas et al.^16^ surveyed 8,495 adults living in Greece using a web-based version of the Active-Q questionnaire (translated to Greek) to compare Physical Activity (PA) before and during lockdown. The change in overall PA (measured in MET-min/week) from before to during lockdown was significant, at −16.3% (95% CI, −17.3 to −15.4). Specifically, this was while in daily occupational, transportation, and sporting activities, where it was −52.9%, −41.1% and −23.9%, respectively. This highlighted that the lockdown period and working from home in Greece was highly associated with a negative change in overall PA. These results are robust despite the likely selection bias associated with the web-based method of data collection and the fact that 22% of the Greek population do not have home internet access. The fact that the survey respondents were all under age 49 years also contributes to the selection bias. Despite these limitation we classified this before-after study as category 1.

Katewongsa, et al. used data from an annual survey on physical activity (Thailand’s Surveillance on Physical Activity (SPA), which is based on the Global Physical Activity Questionnaire version 2.0) in 2019 and 2020 to examine the effect of the COVID-19 pandemic on moderate-to-vigorous physical activity (MVPA) of Thai adults and assessed the effects of the national curfew policy and health promotion campaigns in influencing physical activity during the pandemic. MVPA among Thai adults decreased by 20% (from 74.6% in 2019 to 54.7%) in 2020. There was a substantial reduction in the cumulative minutes of MVPA of Thai adults before and during the COVID-19 pandemic (from 580 to 420, t = 11.864, p < 0.001). Limitations of this study include the different methods of data collection pre- and during the pandemic (face-to-face interviews vs. online self-reporting). There is also a high-risk of selection bias as PA levels in the elderly could not be document.

*Low physical activity summary. There is a clear impact on PA from lockdowns and restrictions – that is likely not completely or even largely due to confounding by the pandemic per se (although there was no category 1a evidence). Based on the studies we included in Category 1b (and therefore Figure 1 in the main paper), there is a 7% to 40%reduction in PA (and presumably METS). This could be approximated (on a log scale) as a ratio of 0.75 (95% UI 0.6 to 0.93., i.e. anchored to lowest and highest estimate in Figure 2).*

##### Tobacco smoking

Three studies investigating smoking behaviour met the inclusion criteria – but only two measures of interest (both prevalence) were of high enough internal validity to make category 1b. No category 1a estimates were found.

Jackson et al (2020)^17^ used a before/after design to examine changes in cigarette smoking following the first lockdown in the UK. The analysis was based on data from the Smoking and Alcohol Toolkit Study (a monthly cross-sectional surveys of adults (≥16 years) in England (Beard et al. 2015, Jackson et al 2018). Due to the lockdown, the March 2020 survey did not occur, and the April 2020 survey was conducted by telephone (whereas pre-lock April 2019 to February 2020 surveys were face-to-face). The smoking prevalence in April 2020 was 17.0% (95% 15.2–18.9) compared to 15.9% (15.4–16.4) for the previous year, a difference of 1.1% point (simulated 95% CI -0.8% to 3.1%) or a relative difference of 7.0% (simulated 95% CI -5.1% to 19.4%).

Jackson et al^17^ also estimate smoking cessation in April 2020 (after lockdown) compared to April 2019 to February 2020 data. There were differences in quit attempts and successful quitting – but this was self-reported for the *last year* and was thus not specific to the month post-lockdown, and so this study outcome was rejected from our category 1. It is notable that due to the lockdown, data collection shifted to telephone-based interview compared to face-to-face in the 11-month before comparison period – suggestion response bias may be at play for the observed differences of a 39.6% versus 29.1% reporting of quit attempts (adjusted odds ratio 1.56, 95% CI 1.23 to 1.98) and a 21.3% versus 13.9% quitting success (OR 2.01, 95% CI 1.22 to 3.33) and 8.8% versus 4.1% cessation (OR 2.63, 95% CI 1.69 to 4.09).

Using the UK Household Longitudinal Study Niedwiedz et al (2020)^2^ determined differences in smoking prevalence in three historic waves centred on 2016, 2017 and 2018 respectively, with data collected April 2020 during a lockdown.^18^ They use a multilevel model with survey year included to capture time trends, and estimate a relative percentage decline in smoking of 11.0% (95% CI 3% to 18%; derived from RR in their Table 2).

Westrupp et al (2020)^19^ surveyed 2,365 Australian parents (81% mothers) with a child under age 18 years to examine the effects of the pandemic on parents and children through a series of psychosocial and health behaviour outcomes, including parent smoking behaviour. The authors compared baseline outcome estimates to pre-pandemic Australian datasets. For the smoking outcome, the survey findings were matched to data from the Longitudinal Study of Australian Children – but the sampling frames (paid and unpaid responders to internet advertising during lockdown, versus population representative sampling in LSAC) render the comparisons biased (7.7% versus 18.6% in pre-pandemic data) and not included in our category 1.

*Smoking prevalence summary. The two studies in category 1 showed modest effects, in opposing direction. Therefore, we conclude that there is moderate evidence of no change in smoking prevalence. For example, it would be reasonable to assume a best estimate of 0% change in current smoking prevalence, but with +/- 5% SD … pending further evidence that either or both shifts the best estimate and narrows uncertainty. Even more uncertain is if there is any impact of lockdowns on smoking prevalence, whether it persists after lockdowns finish.*

*However, we note that smoking is driven by changes in initiation and cessation; better studies are required on the impact of lockdowns on cessation rates – and if any effect is found, whether it persists beyond lockdowns.*

Table S.2 provides further details on the included studies reporting on the risk factors considered

Table S.3: Risk factor studies

| **Risk factor / First author^(ref)^,** | **Title** | | **Country/ population** | | **Design** | **Published effect sizes** |
| --- | --- | --- | --- | --- | --- | --- |
| Alcohol consumption | | | | | | |
| Colbert et al.^3^ | COVID‐19 and alcohol in Australia: Industry changes and public health impacts | | Australia | | Comparing two time periods, before and during lockdown.  Stage 0 vs 3 | The study found a 34% increase in alcohol spending in March 2020, compared to 2019. The following week showed a 28% increase compared with 2019. |
| Tran et al.^4^ | Alcohol use and mental health status during the first months of COVID-19 pandemic in Australia | | Australia | | Online survey comparing pandemic with pre pandemic  A total of 13,829 people contributed complete data and were included in the analysis. | 3253 (20.9%) stated they drink ‘more than I used to’ 95% CI (19.7; 22.1)  1185 (10.5%) drink  ‘Less than I used to’ 95% CI (9.5; 11.6)  6158 (43.9%) ‘About the same’ [42.4; 45.4]  3233 (24.7%) Don't drink alcohol [23.3; 26.1] |
| Niedzwiedz et al.^2^ | Mental health and health behaviours before and during the initial phase of the COVID-19 lockdown: longitudinal analyses of the UK Household Longitudinal Study | | Australia | | Survey, containing results from more than 27 000 people, comparing data before and during lockdown,  Stage 0 vs 3 | the proportion of people drinking four or more times per week increased (RR=1.4, 95% CI 1.3 to 1.5), as did binge drinking (RR=1.5, 95% CI 1.3 to 1.7). |
| Rolland et al.^5^ | Global Changes and Factors of Increase in Caloric/Salty Food Intake, Screen Use, and Substance Use During the Early COVID-19 Containment Phase in the General Population in France: Survey Study | | France | | Wed based survey during containment in March (day 8 to 13) in France.  Stage 0 vs 3 | Regarding alcohol use, 7108/11,391 (62.40%) respondents were found to use alcohol more or less regularly. Among them, 4109/7108 (57.82%) reported that they had not changed their average daily use of alcohol, whereas 1654 (23.27%) moderately increased their alcohol use, 107 (1.50%) increased their alcohol use in a difficult-to-control manner, 1167 (16.4%) declared having reduced or stopped without craving/withdrawal, and 70 (0.98%) having reduced with craving/withdrawal. |
| Dumas et al.^6^ | What Does Adolescent Substance Use Look Like During the COVID-19 Pandemic? Examining Changes in Frequency, Social Contexts, and Pandemic-Related Predictors | | Canada | | Online survey comparing 3 weeks before and after lockdown was in place  Stage 0 vs 3 | The percentage of adolescents who used alcohol did not change significantly from pre- COVID to post-COVID (28.6%–30.1%, p = .23). In contrast, the frequency of alcohol use (i.e., average number of alcohol-using days) increased significantly (F(1, 1,029) = 5.23, p = .02). |
| Sidor et al.^20^ | Dietary Choices and Habits during COVID-19 Lockdown: Experience from Poland | | Poland | | Cross-sectional online survey (n = 1097) before and during lockdown  Stage 0 vs 3 | Over 43.0% and nearly 52% self-reported eating and snacking more, respectively, and these tendencies were more frequent in overweight and obese individuals. Almost 30% and over 18% experienced weight gain (mean ± SD 3.0 ± 1.6 kg) and loss (-2.9 ± 1.5 kg), respectively. |
| Kilian et al.^21^ | [Alcohol consumption during the COVID-19 pandemic in Europe: a large-scale cross-sectional study in 21 countries (preprint](https://www.researchgate.net/publication/348585780_Alcohol_consumption_during_the_COVID-19_pandemic_in_Europe_a_large-scale_cross-sectional_study_in_21_countries_preprint)) | | European Union | | Cross-sectional online survey with 31 964 people conducted between April 24 and July 22 of 2020 | Across all countries, the consumption-change score indicated an average decrease of 0.14 (95% CI: -0.18, -0.10; p < .001). The average consumption-change score ranged between –0.37 (95% CI: -0.52, -0.22; p < .001) in Albania to +0.10 (95% CI: 0.03, 0.17; p = .004) in the UK. With regard to overall consumption change, almost half of the respondents with a negative consumption-change score (decrease level 1 to 6) reported to have substantially reduced their consumption (5,967 of 12,709 respondents with decrease level ≥ 3). This in contrast to drinkers with a positive consumption-change score (increase level 1 to 6), who seldomly reported substantial increases (22% or 1,568 of 7,240 respondents with increase level ≥ 3). |
| **Body mass index (BMI)** | |  | |  | |  |
| Pellegrini et al.^7^ | Changes in Weight and Nutritional Habits in Adults with Obesity during the “Lockdown” Period Caused by the COVID-19 Virus Emergency | | Italy | | Survey of those attending an obesity clinic after 1 month of enforced lockdown.  Stage 0 vs 3 | On average, during the lockdown period, self-reported weight and BMI significantly increased by 1.51 kg (p < 0.001) and 0.58 kg/m2 (p < 0.001), respectively. |
| He et al. ^8^ | Changes in Body Weight, Physical Activity, and Lifestyle During the Semi-lockdown Period After the Outbreak of COVID-19 in China: An Online Survey  Category 2 | | China | | Online Survey during lockdown analysing habits before and during lockdown  Stage 0 vs 3 | Both females and males with BMI < 24 gained weight (before vs after) - female: 51.1 ± 4.1 vs 53.3 ± 5.9 kg, P<0.001; male: 65.6 ± 5.8 vs 67.3 ± 5.5 kg, P<0.001  Males with BMI ≥ 24 lost weight and females with BMI ≥ 24 gained weight (before vs after 62.9 ± 4.2 vs 63.8 ± 5.4 kg, P = 0.042 and 77.3 ± 7.5 vs 76.4 ± 6.9 kg, P = 0.003 for female and male, respectively). The change in body weight by BMI category during the semi-lockdown period was significant for males and females (p<0.001 vs p=0.06). |
| Yang et al.^22^ | Obesity and activity patterns before and during COVID-19 lockdown among youths in China  Category 1 | | China | | Survey to assess changes in obesity and activity patterns before, and during Chinas hard lockdown  Stage 0 vs 3 | BMI increased in overall youth (21.8-22.6, P < .001) and in all subgroups: high school (22.7-23.8, P < .001), undergraduate (21.4 to 22.2, P < .001), and graduate students (21.4-22.3, P < .001). " |
| **Intimate partner violence (IPV)** | |  | |  | |  |
| Beigelman, et al.^9^ | COVID-19 and help-seeking behaviour for intimate partner violence victims | | Spain | | Rates of emergency calls during each month of the lockdown, compared with previous years. Calls then compared with mobility. | An increase of 41 percentage points compared to pre pandemic levels. Using detailed mobile phone data to measure mobility levels, we document stronger effects in provinces whose effective mobility reduction was more intense. |
| Lundin et al.^10^ | Gender-based violence during the COVID-19 pandemic response in Italy | | Italie | | Data compared though months of pandemic response, and to previous years. | Access to anti violence services increased 191% (1306 in previous years (2016-18) to 2496 in the same period in 2020). |
| **Physical activity** | | | | | | |
| Gallo et al.^11^ | The Impact of Isolation Measures Due to COVID-19 on Energy Intake and Physical Activity Levels in Australian University Students | | Australia | | Survey (Active Australia Survey) comparing two | In males, comparing year 2020 to 2018/19 combined had significant reduction in walking participation but no difference for vigorous activity. Females had a reduction for walking participation in 2020 compared with 2018/19 (chi2 6.30, p<0.05) but no difference for vigorous activity. |
| Tison et al.^14^ | Worldwide effect of COVID-19 on physical activity: a descriptive study | | World* | | Descriptive study of literature surrounding step count changes  Stage 0 vs 3 | Worldwide, within 10 days of the pandemic declaration, there was a 5.5% decrease in mean steps (287 steps), and within 30 days, there was a 27.3% decrease in mean steps (1432 steps). Wide regional variation in average step count change and in the timing and rate of that change e.g. a 48.7% maximal decrease after a nationwide lockdown in Italy whereas Sweden, to date, has primarily advocated for social distancing and limitations on gatherings had a 6.9% maximal decrease. |
| López-Sánchez et al.^23^ | Comparison of physical activity levels in Spanish adults with chronic conditions before and during COVID-19 quarantine | | Spain | | Survey comparing PA rates before and during lockdown | Moderate-intensity PA significantly decreased in Spanish people with chronic conditions during quarantine (mean difference: 30 min/day; P < 0.001). The decrease was significant in both males (mean difference: 22.1 min/day; P = 0.006) and females (mean difference: 33.2 min/day; P < 0.001).   Vigorous-intensity PA significantly decreased in Spanish males with chronic conditions during COVID-19 quarantine [mean difference: 15.5 min/day; P = 0.025].  Moderate-intensity PA significantly decreased during quarantine in those diagnosed with asthma (mean difference: 26.2 min/day; P = 0.026), hypercholesterolaemia (mean difference: 39.0 min/day; P = 0.011), chronic skin disease (mean difference: 44.3 min/day; P = 0.004) and haemorrhoids (mean difference: 50.1 min/day; P = 0.009; table 3). No significant differences in vigorous-intensity PA by chronic condition before and during quarantine. |
| Luciano et al.^15^ | COVID-19 lockdown: physical activity, sedentary behaviour and sleep in Italian medical students  Category 2 | | Italy | | Survey determining IPAQ score before and during lockdown | A median IPAQ score of 1588 METmin/week in 6th year students before lockdown and 960 MET-min/week in 6th year students during lockdown, with a reduction of 628 MET-min/week (p<0.01). In 6th year students, time dedicated to moderate and vigorous activity increased during lockdown (p<0.01 for moderate physical activity, p=0.30 for vigorous physical activity), while walking time decreased (p<0.01). Furthermore, an increase in sedentary behaviour was observed: 6th year students spent a median of 8 hours/day sitting before lockdown, and 10 hours/day during lockdown (p<0.01). |
| Bourdas et al.^16^ | Impact of COVID-19 Lockdown on Physical Activity in a Sample of Greek Adults | | Greece | | Web-based survey determining IPAQ score before and during lockdown | For each PA, duration(min/week), the EE scored in the METs was calculated (e.g., PA of 4 METs × 30 min/day × 2 times per week = 240 MET-min/week).   The relative frequencies of overall sporting or exercise activities of participants who PRE had experienced said activity at least once per month were significantly reduced by 8.6% (95%CI, 7.9–9.3) during the quarantine period.  The relative frequency of overall participation in competitive sports of 16.9% (95%CI, 15.0–18.9) in the PRE condition nearly reached a nadir in the middle of April (2.6% (95%CI, 0.5–4.7)).  The daily energy expenditure (EE) was significantly reduced in the POST condition. The frequency of inactivity significantly increased by 40.6% (95%CI, 38.3–42.9) in the POST condition, while the frequency of moderate and high PA was significantly reduced, by 12.6% (95%CI, 10.5–14.7) and 13.0% (95%CI, 12.0–14.0), respectively. |
| Katewongsa et al.^24^ | The effects of the COVID-19 pandemic on the physical activity of the Thai population: Evidence from Thailand’s Surveillance on Physical Activity 2020 | | Thailand | | Survey (face-to-face interviews for 2019 and online questionnaire for 2020) | The prevalence of sufficient MVPA among Thai adults decreased from 74.6% in Surveillance on Physical Activity (SPA) 2019 to 54.7% in SPA 2020. Males were 1.3 times more likely to have sufficient MVPA during the pandemic compared to females.  Significant difference in the cumulative minutes of MVPA of Thai adults before and during the COVID-19 pandemic (from 580 to 420, t = 11.864, p < 0.001).  The F-test analysis also indicates that a significant difference in PA exists in 2 of 3 periods of the pandemic: Before Curfew (BC) vs. DC, and BC vs. After maximum curfew was relaxed (AC) (F = 70.610, p < 0.001). |
| **Tobacco smoking** | | | | | | |
| Jackson et al.^17^ | Association of the Covid‐19 lockdown with smoking, drinking, and attempts to quit in England: an analysis of 2019‐2020 data | | United Kingdom | | Before-after analysis using monthly cross sectional surveys | Lockdown was not associated with an increase in smoking prevalence (17% vs 15.9%), but was associated with increased quit attempts, quit successes and cessation. |
| Niedzwiedz et al.^18^ | Mental health and health behaviours before and during the initial phase of the COVID-19 lockdown: longitudinal analyses of the UK Household Longitudinal Study | | United Kingdom | | Repeated cross sectional and longitudinal analysis of UK Household Longitudinal Survey. Cross-sectional prevalence estimates calculated with Poisson regression. | Cigarette smoking decreased during lockdown. This was more apparent in younger age groups. Effects seems to be driven by a decline in the number of lighter smokers. Relative risk of smoking was 0.9 (95% CI: 0.8-1.0) |
| Westrupp et al^19^. | Child, parent, and family mental health and functioning in Australia during COVID-19: Comparison to pre-pandemic data | | Australia | | Self-report survey during stage 3 of lockdown. Data compared to pre pandemic data from Australian population-based cohorts | Matched weighted data showed a 3.6% decrease in smoking during the lockdown compared with pre pandemic. |

† Effect measure calculated by authors; CI generated with various simplifying assumptions (e.g. Assuming no covariance between estimates)

#### Health seeking behaviour: cancer screening

No studies met our inclusion criteria for this domain. The vast majority studies identified by our literature search reported specifically on the disruption caused by the pandemic to cancer screening and treatment services. The search also picked up a modelling studies on the likely consequences of pandemic related disruptions. Box 2 in the main manuscript summarises the findings on those studies.

#### Conditions and diseases

##### Anxiety and depression

Four studies met our category inclusion criteria, with five measures of interest.

Papandreou et al (2021)^25^ undertook surveys in Spain and Greece in April/May to June 2020. Assuming their multivariable analyses renders Spanish and Greek respondents exchangeable other than the level of restrictions (which we estimate as an average of Stage 3 in Spain and 1 in Greece based on authors statement from the authors, a difference of 2 stage units), this study provides estimates of depression and anxiety impacts from social restrictions. The regression adjusted difference in depression PHQ-9 scores was 0.21 units (95% CI -0.27 to 0.70) higher in Spain, and the starting mean and SD of 5 and 4.8. PHQ-9 scores are generally log normal, and a score of 10 or more equates to moderate depression. Using these assumptions, and assuming the SD on log scale does not change with lockdown (just the mean), then we can estimate that the tougher restrictions in Spain increased the relative prevalence of depression by 9.6% (95% CI -10.4% to 39.5%). Finally, we therefore estimate that this study implies a 16% (95% CI -17% to 63%) change in depression for Stage 3 compared to Stage 0.

Papandreou et al (2021)^25^ also estimate anxiety differences. Using the same logic and similar assumptions to above, we estimate Spain increased the relative prevalence of anxiety by 52.5% (95% CI 16.5% to 109.5%). Transposing this to a stage 0 to 3 Stage difference, this is a 88% increase in anxiety (28% to 182%). It must be emphasized these calculations are contingent on strong assumptions implicit in Papandreou et al (2021), plus strong assumptions we imposed to generate our metrics of interest.

Foa et al (2021) ^26^provide a useful time series study of mood and affectivity in the UK. The items do not correspond to depression or anxiety per se as clinical diagnoses (e.g. happiness, optimism), but nevertheless the analysis whilst demonstrating falls in affect associated with lockdown also reveals that it is as much – if not more – due to the pandemic per se (i.e. infection rates in the community), with affect improving quite rapidly into lock downs.

Sibley et al. (2020)^27^ use the New Zealand Adult Values Survey to determine changes in psychological distress from pre- to post-lockdown, using the short 6-item Kessler scale – a screening tool for depression and anxiety as one construct. They use two analyses to adjust for confounding: propensity score matching of post-lockdown respondents to pre-lockdown respondents, and within-individual changes over time for those respondents with measures both before and after lockdown. The propensity score analysis found a 0.08-unit (12% of SD) increase in score (95% CI 0 to 0.15), and the within-individual analysis a 0.04-unit (6% of SD) increase (95% CI 0 to 0.07). Categorical analyses splitting the sample by ‘clinical thresholds’ was mixed: mild or moderate distress increased from 16.2% to 21.1%, but serious distress did not change 6.6% to 5.8% with Chi-squared test borderline (p=0.02), This study provides evidence of modest increases in psychological distress, but its use of a short-form screening tool precludes more definitive comment on changes in depression and anxiety as clinical states.

Fancourt et al (2020^28^) analysed within person changes over time in anxiety and depression during the UK initial lockdown, using the GAD-7 and PHQ-9. Unfortunately, it did not include observations outside of lockdown, and therefore there is no variation in exposure. It does however find that anxiety elves waned during lockdown, and that depression levels were constant.

##### Chronic obstructive pulmonary disease (COPD)

Gonzalez et al. investigated the effects of the lockdown put in place in Spain on COPD exacerbation, symptoms and healthcare costs. Data from 310 COPD patients attended a specialised pulmonary consultation clinic in 2019 was collected retrospectively from electronic medical records. A phone interview was conducted to collect data for 2020. Data on exacerbations and symptoms before the pandemic (1^st^ of March to 31^st^ of May 2019) were compared to data collected during the lockdown (1^st^ of March to 31^st^ of May 2019). A 62% decrease in the number of COPD exacerbations was observed. Despite the before-after design, this study was classified as category 2 because it is highly likely that the reported effect of lockdown is heavily confounded by the pandemic itself. For example, through people’s fear of attending a hospital during a pandemic. The study was therefore classified as category 2.

##### Falls

Jacob et al (2020)^29^ investigated the effects of COVID-19-related restrictions and lockdown on severe trauma admissions in an Australian emergency department in New South Wales using a before-after study design. The authors reported a 13-29% decrease in the average number of admissions due to injuries from falls in March/April 2020 compared to the previous three years.

##### Ischaemic Heart Disease and Heart Failure

Five studies were found exploring IHD or heart failures of several types, with all studies presenting rates of health seeking, such as hospitalizations. Unfortunately, no studies analysing incidence, prevalence or mortality of this condition were found, thus there were no category 1 studies.

Andersson et al (2020)^30^ analysed the incidence of heart failure before and after the lockdown in Denmark, using a difference in difference study of hospital records. The authors used a proxy of the weeks leading into lockdown for their study. They found a 44% reduction of heart failure rates during the Danish lockdown, and a 36% decrease in worsening heart failure rates during the lockdown. Caution is advised when interpreting these results as a reduction in actual incidence, as reluctance to enter hospitals may be responsible for this trend.

Similarly, Oikonomou et al (2020)^31^ explore hospital admission trends, to determine the relationship between cardiac disease admissions and restrictive measures for adults in the Greek population, through an observational study compared to data from 2019. They found that cardiology emergency department visits decreased by 53%(p<0.001) during the lockdown, compared to pre-lockdown rates. For reference there was no difference found between 2020 pre-lockdown rates and the 2019 rates of cardiology emergency department visits.

Schiele et al also performed an observational study with hospital admissions data, measuring myocardial infarction in French adults. Admissions decreased by 30% during the lockdown period, with the incidence ratio of 0.69 (95% CI: 0.51-0.70). The authors found no association between this decrease and patient characteristics or regional prevalence of COVID-19. Similarly, Lantelme et al performed a cross sectional, retrospective study of myocardial infarction hospital presentations pre- and post-lockdown in Lyon, France. A 31% decrease in hospitalisations was found for myocardial infarction presentations during lockdown.

Finally, Ball et al (2020)^32^ performed a retrospective, cross sectional study of hospital admissions in the UK, inclusive of pre- and post-lockdown. A 40.2% decrease in cardiac related hospitalisations was found during the lockdown, was found in the UK.

Overall, all five studies found use health resource usage, though slightly different metrics, in describing incidence the change in heart related illnesses during lockdowns. Great caution must be advised when interpreting these results. While all studies found decreases in hospital usage for heart related conditions, there is no evidence to suggest population incidence rates decreased during the lockdown.

##### Interpersonal Violence

Two studies were identified for interpersonal violence using health utilisation outcome measurements. As above, Jacob et al (2020)^29^ also explored trauma presentations at the same Sydney hospital before and after the lockdown. Whilst this study did show that lockdown was associated with fewer overall trauma admissions by 23-34%, there was no evidence of difference in the trauma admissions from assault. Thus, they showed no change for a health utilisation measure of interpersonal violence.

The second, a hospital admission review study from the UK analysed penetrating injuries during the 6-week lockdown. Olding et al^33^, found that penetrating trauma injuries fell by 35% compared to previous years.

##### Road transport injuries

Three studies met the inclusion criteria for road traffic accidents, comparing accident statistics from time periods before, to during lockdown.

Jacob et al (2020)^29^ conducted a before-after study using admission data from a single NSW hospital emergency department in New South Wales (Australia). The authors found that in March/April 2020, there was a 40-52% decrease in the average number of admission due to road traffic collision compared to the same period in over 2016-2019.

Using data from the Missouri State wide Traffic Accident Records System for the first 5 months of 2020*,* Quaraishi et al. (2020)^34^ conducted an interrupted time series analysis to evaluate the effect of mandated lockdown on the traffic accidents. The authors found that the lockdown had no effect on the rates of road traffic accidents resulting in serious or fatal injuries(mean 3.4 vs. 3.7, p=0.42) , but lead to a reduction on those causing no, or non-serious injuries (mean 14.5 vs. 10.8, p<0.0001).

Analysing data from a hospital providing emergency medical services to the North of Dublin (Ireland), Fahy et al^35^, conducted a before-after study comparing the number of trauma admissions during the government month-long lockdown imposed on the 27^th^ of March 2020 to the number of trauma admissions during the same month in 2019. The authors reported an overall 22% reduction in trauma admissions in 2020 and a 60% reduction in the number of patients admitted due to injuries during traffic accidents – 10 in 2019 compared to 4 in 2020. But given small numbers, statistical imprecision is too high to make any conclusion.

##### Self-harm

Four studies on self-harm were found, with hospital admissions studies being the most prevalent outcome metric. Two of these studies found increases in self-harm admissions during lockdowns, with one showing no difference.

Joyce et al^36^ examined hospital admissions at a tertiary emergency department in Christchurch, New Zealand during their strict lockdown. They showed that the proportions of presentations to the emergency department increased during the lockdown increased by 3.5% (95% CI: 0.01%-0.01%).

Similarly, Henry et al.^37^ performed an observational study on hospital before and during lockdown in the UK. This study found an increase in deliberate self-harm presentations within this hospital by 8.8% (no CI given).

One Australian study was sought to determine whether social restrictions and lockdown increased the amount of trauma admissions at a hospital in Sydney. Jacob et al^29^ found no significant increase in self-harm trauma presentations, however, did have very low numbers of patient admissions during their observation period (n=8).

##### Stroke

7 studies were found exploring health service utilisation for those who have had a stroke, but no studies were found around prevalence, mortality, or morbidity.

Using data from 4 stoke centres in Germany, Ikenberg et al.(2020)^38^ found no differences in daily referrals in the first 15 weeks of 2020, compared to the same period in 2019. Similar portions of daily referred patients were categorized as stroke during the first 4 weeks of 2020 (pre-lockdown) (50% (IQR 13-100)) and lockdown (50% (IQR 31-67)). There were no differences in daily referral numbers of ischemic strokes, hemorrhagic stroke, and transient ischemic attacks between both periods. However, stroke severity as measured by the National Institutes of Health Stroke Scale (median 3 (IQR 0-7) versus 6 (IQR 1-15.5) points; p = 0.04) increased during the lockdown.

In a similar design, Schlachetzki et al (2020)^39^ evaluated the effect of the COVID-19 lockdown on stroke consultations and treatment recommendations using the stroke consultant database of 12 hospitals within a telestroke network in Germany. Upon lockdown in mid-March 2020, a reduction in recommendations for recombinant tissue plasminogen activator (rtPA) for acute ischemic stroke was observed, compared to the preceding three years (14.7% [2017–2019] vs. 9.2% [2020], p¼0.0232). Recommendations for stroke treatments such as endovascular treatment (EVT) were higher in January to mid-March 2020 compared to 2017–2019 (5.4% [2017–2019] vs. 9.3% [2020],p¼0.0013) highlighting its increasing importance. Following the lockdown in mid-March 2020 the number of EVT decreased back to levels in 2017–2019 (7.4% [2017–2019] vs. 7.6% [2020], p¼0.1719). Absolute numbers of ischemic stroke decreased in parallel to mobility data.

In a retrospective analysis of hospital admissions, Hoyer et al (2020)^40^ found a significant reduction in the number of admissions for stroke related events by 35.9% (p = 0.005). In addition, significantly more patients arrived by ambulance during the COVID-19 lockdown (2019: 75.7%, 2020: 94.2%; p = 0.001), suggesting people did not present in a timely manner, leaving their condition to deteriorate before calling emergency services.

Frisullo et al (2020)^41^ found that during the Italian the lockdown, a significant increase in time between onset and admission (median = 387 vs 161 min, p = 0.001) was observed, along with a significant reduction of the total number of thrombolysis (7 vs 13, p = 0.033), a non-significant increase of thrombectomy (15 vs 9, p = 0.451), and a significant increase in door-to-groin time (median = 120 vs 93 min, p = 0.048). No relevant difference was observed between 2019 and 2020 in the total number of patients admitted.

A similar study by Pailiwal et al (2020)^42^ in a stroke centre in Singapore, discovered the number of stroke activations presented significant decline (p = 0.004, 95% CI 6.513 to − 2.287), as the number of COVID-19 cases increased, whilst proportion of activations receiving acute ischemic stroke therapy remained stable (p = 0.519, 95% CI − 1.605 to 2.702).

Kristoffersen et al conducted two separate studies on stroke admissions during the lockdown from a single hospital dataset in Norway, due to concerns that public anxiety around COVID‐19 discourages patients from seeking medical help.

Kristoffersen et al (2021)^43^ found a decrease in weekly stroke admissions during lockdown 2020 to the preceding 5 years. There was an average of 21.4 (SD 4.7) before to 15.0 (SD 4.2) during and 17.2 (SD 3.3) after (p < 0.011). The proportion of mild ischemic and haemorrhagic strokes was also lower during lockdown with 66% before, 57% during and 68% after (p = 0.011).

Kristoffersen et al (2021)^43^ found a decrease in weekly stroke admissions during lockdown 2020 to the preceding 5 years. There was an average of 21.4 (SD 4.7) before to 15.0 (SD 4.2) during and 17.2 (SD 3.3) after (p < 0.011). The proportion of mild ischemic and haemorrhagic strokes was also lower during lockdown with 66% before, 57% during and 68% after (p = 0.011).	Kristoffersen et al (2020)^44^ found there were 21.8 (SD 4.7) admissions weekly before the lockdown and 15.0 (SD 4.2) admissions weekly during the lockdown (P.008), and found a 32% reduction in the weekly number of admissions for stroke (29%) and TIAs (41%) during the first seven weeks of the pandemic lockdown in Norway. In the multivariable logistic regression model for ischemic stroke (adjusted for sex, age, living alone and NIHSS ≤ 5), there was an increased OR of 2.05 (95% CI 1.10‐3.83, P = .024) for not reaching hospital within 4.5 hours during the lockdown as compared to the period before the lockdown.

##### Suicide

One study met our inclusion criteria for suicide. Leske et al (2020)^45^ analysed data from the Queensland Suicide Register (iQSR), a state-wide real-time suicide surveillance system, to determine whether Queensland's COVID-19 Public Health Emergency Declaration, announced on Jan 29, 2020, and the movement restrictions announced in the subsequent days, affected suspected suicides from Feb 1 to Aug 31, 2020, compared with the preceding 5 years. 14·85 deaths per 100 000 people was recorded before the declaration, and 14·07 deaths per 100 000 people afterwards. An interrupted time-series Poisson regression model unadjusted (rate ratio [RR] 0·94, 95% CI 0·82–1·06) and adjusted for overdispersion, seasonality, and pre-exposure trends (RR 1·02, 95% CI 0·83–1·25) indicated no evidence of a change in suspected suicide rates. Additionally, there were no increases in the motives for suspected suicides, including recent unemployment, financial problems, relationship breakdown, or domestic violence from February to August, 2020, compared with the pre-exposure period.

*Suicide summary. We found no reliable evidence of suicide rate changes in response to lockdowns and restrictions.*

Table S.3 provides further details on the included studies reporting on the diseases and conditions considered.

Table S.4: Disease and condition studies

| **Diseases & injuries** | **Title** | | **Country** | **Design** | | **Effect measures as reported by authors** |
| --- | --- | --- | --- | --- | --- | --- |
| **Anxiety and Depression** | | | | | | |
| Papandreou, et al.^25^ | Comparing eating behaviours, and symptoms of depression and anxiety between Spain and Greece during the COVID‐19 outbreak: Cross‐sectional analysis of two different confinement strategies | | Spain and Greece | Web‐based cross‐sectional survey. Comparing the Spain lockdown (stage 4) vs Greek lockdown (stage 3), with results after the lockdown ended | | Proportion of respondents with moderate to severe anxiety symptoms was 13.6% in Spain, vs 18.8% in Greece. |
| Fancourt et al.^28^ | Trajectories of depression and anxiety during enforced isolation due to COVID-19: longitudinal analysis of 59,318 adults in the UK with and without diagnosed mental illness | | United Kingdom | Weekly panel data during the lockdown, and then as restrictions were eased.  Potentially stage 3 vs 2 vs 1**.** | | No pre-and post-lockdown values given. |
| Papandreou et al.^28^ | Comparing eating behaviours, and symptoms of depression and anxiety between Spain and Greece during the COVID‐19 outbreak: Cross‐sectional analysis of two different confinement strategies, with Spain averaging about Stage 3 | | Spain and Greece | Web‐based cross‐sectional survey. Comparing the Spain lockdown (stage 4) vs Greek lockdown (stage 0), with results after the lockdown ended | | Patient health questionnaire responses were 5.0 (Spain) vs 5.6 (Greece). In multivariable linear regression comparing the two countries, however, this difference was non-significant (0.21 units on PHQ-9, 95% CI -0.27 to 0.70). Expressed as percentage change, ASSUMING multivariable adjustment has made Greece exchangeable counterfactually for Spain, and using 5.0 as baseline for spain, this is |
| Foa et, al^26^ | COVID-19 and Subjective Well-Being: Separating the Effects of Lockdowns from the Pandemic  Category 2 | | United Kingdom | Weekly cross-sectional survey of 1,890-2,071 respondents in Great Britain before and during lockdown  Stage 0 vs 3 | | The one-month period in lockdown was associated with reduced negative affect of around -9% relative to a pre-pandemic baseline, rising to -17% when the sample space is restricted to the period following lockdown onset. However, a composite index demonstrated the largest falls in affect occurred just before lockdown, and then rose in the first couple of weeks of lockdown. |
| Sibley et al.^27^ | Effects of the COVID-19 pandemic and nationwide lockdown on trust, attitudes toward government and well-being | | New Zealand | Matched sample survey before, during and after lockdown  Stage 0 vs 3 | | Not significant |
| Lei Lei et al. ^46^ | Comparison of prevalence and associated factors of anxiety and depression among people affected by vs people unaffected by Quarantine during the COVID-19 epidemic in Southwestern China | | China | Matched sample survey of those in quarantine vs not quarantining.  Stage 1 vs 4 | | Prevalence of self-reported depression rose from 11.9% to 22.4% in survey participants who were quarantined. |
| **Chronic obstructive pulmonary disease (COPD)** | | | | | | |
| González et al.^47^ | Clinical consequences of COVID-19 lockdown in patients with  COPD: results of a pre-post study in Spain | | Spain | Review of hospital admissions data before lockdown and phone interview during lockdown  (consisting in the limitation of free movement except for acquiring food and medicines,  seeking for healthcare, and attending work in essential services)  Stage 0 vs 4 | | A 62% decrease in the number of COPD exacerbation was observed – particularly in the number of severe exacerbations (76% reduction in 2020 compared to 2019) |
| **Falls** | | | | | | |
| *Jacob et al.^29^* | Impact of societal restrictions and lockdown on trauma admissions during the COVID-19 pandemic: a single-centre cross sectional observational study | | Australia | Review of hospital admissions data a single‐centre. Cross‐sectional observational study before and during lockdown  Stage 0 vs 3 | | There was a 23–34% decrease (P = 0.018) in the mean monthly average trauma admissions during March/April 2020 compared with previous years 2016–2019. In addition, there was a 13–29% decrease (P = 0.020) in admissions due to falls. |
| **Ischaemic Heart Disease** | | | | | | |
| Andersson et al.^30^ | Incidence of New-Onset and Worsening Heart Failure Before and After the COVID-19 Epidemic lockdown | | Denmark | Difference in difference study of hospital records  (Stage 0 vs 3?) | | In lockdown, rates of new onset HF decreased on 2019 rates (1.26 vs 2.25 per 1000 persons) and decreased for worsening HF (0.63 vs 0.99 per 1000). |
| Oikonomou et al. ^31^ | Hospital attendance and admission trends for cardiac diseases during the COVID-19 outbreak and lockdown in Greece | | Greece | Observational study, comparing to pre pandemic, pandemic pre lockdown, lockdown and post lockdown | | Cardiology ED visits decreased by 53% compared to pre lockdown rates (p<0.001) and no difference between 2019 data and COVID-pre lockdown data (p= 0.48) 2019 data. This was then increased in the post lockdown period (% not given) |
| Mesnier et al.^48^ | Hospital admissions for acute myocardial infarction before and after lockdown according to regional prevalence of COVID-19 and patient profile in France: a registry study | | France | Observational hospital admissions study, pre- and post-lockdown | | Admissions for acute myocardial infarction decreased by 30% during the lockdown period. Incidence ratio 0.69 (95% CI 0.51-0.70). More of an impact from Non- STEMI |
| Ball et al^32^ | Monitoring the indirect impact of COVID-19 pandemic on services for cardiovascular disease in the UK | | United Kingdom | Comparative study of hospital presentations, including pre and during lockdown. Retrospective cross sectional | | 40.2% decrease in Cardiac related hospitalisations during lockdown period. |
| Lantelme, et al.^49^ | Worrying decrease in hospital admissions for myocardial infarction during the COVID-19 pandemic | | France | Comparative study of hospital presentations, including pre and during lockdown, and previous years. Retrospective cross-sectional | | 31.0% decrease in hospitalisations for myocardial infarction |
| **Interpersonal violence (IV)** | | | | | | |
| Olding et al.^33^ | Penetrating trauma during a global pandemic: changing patterns in interpersonal violence, self-harm and domestic violence in the COVID-19 outbreak | | United Kingdom | Review of hospital admissions data before and during lockdown  Stage 0 vs 3 | | Overall penetrating trauma injuries during the lockdown decreased by 35% compared to previous years. |
| Jacob et al.^29^ | Impact of societal restrictions and lockdown on trauma admissions during the COVID-19 pandemic: a single-centre cross sectional observational study | | Australia | Review of hospital admissions data a single‐centre. Cross‐sectional observational study before and during lockdown  Stage 0 vs 3 | | Overall, trauma admissions fell by 23-34% during the lockdown period. However, no significant difference in number of assaults. |
| **Road transport injuries** | | | | | | |
| Jacob et al.^29^ | Impact of societal restrictions and lockdown on trauma admissions during the COVID‐19 pandemic: a single‐centre cross‐sectional observational study | | Australia | Review of hospital admissions data a single‐centre. Cross‐sectional observational study before and during lockdown  Stage 0 vs 3 | | There was a 23–34% decrease (P = 0.018) in the mean monthly average trauma admissions during March/April 2020 compared with previous years 2016–2019. In addition, there was a 40–52% decrease (P = 0.025) in admissions due to road traffic collisions. |
| Qureshi et al.^34^ | Mandated societal lockdown and road traffic accidents | | United States | Review of hospital admissions data before and during lockdown  Stage 0 vs 3 | | The daily counts of road traffic accidents decreased during the entire period. Overall, the total daily number of road traffic accidents varied from 17.9 ± 6.1 (before lockdown) to 14.4 ± 4.6 (during lockdown) and 18.1 ± 6.4 (after lockdown) per day. The daily number of serious or fatal traffic accidents ranged from 3.4 ± 1.8 (before lockdown) to 3.7 ± 2.1 (during lockdown) and 4.4 ± 2.4 (after lockdown). |
| Kamine et al.^50^ | Decrease in Trauma Admissions with COVID-19 Pandemic | | United States | Data comparisons before and during lockdown  Stage 0 vs 3 | | There were significant decreases in overall trauma admissions (57.4% decrease, p<0.001); motor vehicle collisions (MVC) (80.5% decrease, p<0.001); and non-MVCs (45.1% decrease, p<0.001) from February–April 2020. |
| Fahy et al.^35^ | Analysing the variation in volume and nature of trauma presentations during COVID-19 lockdown in Ireland | | Ireland | Comparison of trauma presentations from 27 March 2019 to 27 April 2019 versus 27 March 2020 to 27 April 2020.  Stage 0 vs 3 | |  |
| **Self-harm** | | | | | | |
| Joyce, et al. ^36^ | Mental Health Presentations to Christchurch Hospital Emergency Department During COVID-19 Lockdown | | New Zealand | Review of hospital admissions data before and during lockdown  Stage 0 vs 4 | | The proportion of admissions due to self-harm increased 3.5% compared to previous years. |
| Jacob et al.^29^ | Impact of societal restrictions and lockdown on trauma admissions during the COVID-19 pandemic: a single-centre cross sectional observational study | | Australia | Review of hospital admissions data before and during lockdown  Stage 0 vs 3 | | There was no difference in self harm trauma found during the lockdown period, compared with the previous four years. Sample size too small for p-value. |
| Henry et al. ^37^ | The effect of COVID-19 lockdown on the incidence of deliberate self-harm injuries presenting to the emergency room | | United Kingdom | Observational study, and review of hospital admissions data before and during lockdown  Stage 0 vs 3 | | Deliberate self-harm presentations increased from 103 in 2019 to 113 in 2020 (p<0.001). The percentage of ED visits being self-harm increased from 1.98% to 3.69%. |
| Chen et al. ^51^ | |  | |  |  | |
| **Stroke** |  | |  |  | |  |
| Kristoffersen et al.^44^ | Effect of COVID‐19 pandemic on stroke admission rates in a Norwegian population | | Norway | Analysing admissions data, - January 3 to March 12 was defined as before, and March 13 to April 30 as during lockdown.  Stage 0 vs 3 | | There were 21.8 (SD 4.7, range 29‐14) admissions weekly before the lockdown and 15.0 (SD 4.2, range 21‐8) admissions weekly during the lockdown (t test, P = .008).  In the multivariable logistic regression model for ischemic stroke (adjusted for sex, age, living alone and NIHSS ≤ 5), there was an increased OR of 2.05 (95% CI 1.10‐3.83, P = .024) for not reaching hospital within 4.5 hours during the lockdown as compared to the period before the lockdown. |
| Ikenberg et al.^38^ | Code Stroke Patient Referral by Emergency Medical Services During the Public COVID-19 Pandemic Lockdown | | Germany | Retrospective single-center study at a Bavarian Stroke Center. 1st of January 2020 and the 19th of April 2020 were the study periods. The public lockdown started on 21st of March and ended on 19th April 2020  Stage 0 vs 3 | | A total of 171 patients was referred during pre-lockdown and 70 patients during lockdown. There were no differences in EMS-based daily referral of Code Stroke patients between pre-lockdown and lockdown. Comparable portions of daily referred patients were categorized as stroke mimic during pre-lockdown (50% (IQR 13-100)) and lockdown (50% (IQR 31-67)). By trend, there was a weak association between daily availability of our Stroke Unit and daily admission numbers (Pearson r = 0.16, p = 0.10). |
| Schlachetzk et al.^39^ | Low stroke incidence in the TEMPiStelestroke network during COVID-19pandemic – effect of lockdown onthrombolysis and thrombectomy | | Germany | This study was focused on data collected during the first four months of 2020, which included the emergence of COVID-19 through the first two months of social distancing/region shutdown, compared data collected during the same months in the years 2017–2019. Stage 0 vs 3 | | Upon lockdown in mid-March 2020, we observed a significant reduction in recommendations for rtPA compared to the preceding three years (14.7% [2017–2019] vs. 9.2% [2020], p¼0.0232). |
| Paliwal et al.^42^ | Impact of the COVID‑19 pandemic on hyperacute stroke treatment:  experience from a comprehensive stroke centre in Singapore" | | Singapore | This study compared the situation in the preceding months before and after DORSCON Orange (lockdown) was activated in the current COVID-19 pandemic  Stage 0 vs 4 | | Across the study period, number of stroke activations showed significant decline (p = 0.004, 95% CI 6.513 to − 2.287), as the number of COVID-19 cases increased exponentially, whilst proportion of activations receiving acute recanalization therapy remained stable (p = 0.519, 95% CI − 1.605 to 2.702). |
| Kristoffersen et al.^43^ | Stroke admission rates before, during and after the first phase of the COVID-19 pandemic | | Norway | Analysis of all patients discharged from Akershus University Hospital with a diagnosis of transient ischemic attack (TIA) or acute stroke from January to September 2020 | | A transient decrease in weekly stroke admissions during lockdown were observed, from an average of 21.4 (SD 4.7) before to 15.0 (SD 4.2) during and 17.2 (SD 3.3) after (p < 0.011). The proportion of mild ischemic and haemorrhagic strokes was also lower during lockdown with 66% before, 57% during and 68% after (p = 0.011). |
| Hoyer et al.^40^ | Decreased admissions and change in arrival mode in patients with cerebrovascular events during the first surge of the COVID-19 pandemic | | Germany | Analysis of data of patients presenting with a cerebrovascular event to the emergency department during weeks 12–17/2020 were compared to data from the respective weeks in 2019. | | A significant reduction in the number of admissions by 35.9% (p = 0.005) was observed due the COVID-19 epoch. In addition, significantly more patients arrived by ambulance during the COVID-19 epoch (2019: 75.7%, 2020: 94.2%; p = 0.001) |
| **Suicide** | | | | | | |
| Leske et al.^45^ | Real-time suicide mortality data from police reports in Queensland, Australia, during the COVID-19 pandemic: an interrupted time-series analysis | | Australia | Retrospective data analysis from the QLD Suicide Register (iQSR) a state-wide suicide surveillance system,that uses dara from police and coroner reports. | | 3793 suspected suicides were recorded with an unadjusted monthly rate of 14·85 deaths per 100 000 people (from Jan 1, 2015, to Jan 31, 2020) and 443 suspected suicides were recorded with an unadjusted monthly rate of 14·07 deaths per 100 000 people (Feb 1, 2020, onwards). |

### 3. Literature search strategies and PRISMA charts

##### Tobacco smoking

**Literature search criteria (PICO framework)**

| **Inclusion criteria** | |
| --- | --- |
| Population | People in OECD countries, as well as China and HK, and Singapore. |
| Intervention | COVID 19 management related policies |
| Comparators | Different stages of the policy intervention |
| Outcome | Changes in tobacco consumption |
| Language restrictions | None |
| Search dates | Restricted to literature published between 1 January 2020 and 21 January 2021 |
| **Exclusion criteria** | |
| Population | Non-Human based studies. |
| Study design | Case Studies, surveys with no temporal or spatial comparative group |

**Database Search Terms**

**Pubmed**

| **ID** | **Search** | **Hits** |
| --- | --- | --- |
| #1 | (lockdown* OR quarantine) AND (tobacco) | 38 |
| #2 | (lockdown* OR quarantine OR Covid policy) AND (tobacco) | 72 |
| #3 | (tobacco OR smoking) AND (Australia) AND (Covid) | 47 |
| #4 | (lockdown* OR quarantine OR stay-at-home) AND (tobacco) | 43 |
| #5 | (lockdown* OR stay-at-home OR stay at home) AND (tobacco OR smoking) | 94 |
| #6 | (lockdown* OR quarantine OR Covid policy) AND (tobacco) AND Australia | 3 |
| #7 | **(lockdown* OR quarantine OR Covid policy) AND (smoking OR tobacco)** | 176 |

**Google Scholar**

Search date 22^nd^ Jan 2021. Restricted to articles published since 1 January 2020 and sorted by relevance. The first ten pages of each search was assessed due to the large number of hits.

| **ID** | **Search** | **Hits** |
| --- | --- | --- |
| #1 | lockdown* OR stay-at-home OR stay at home AND tobacco OR smoking AND Australia | 1400 |
| #2 | Smoking rates during Australian lockdown | 1270 |
| #3 | Tobacco use Australian lockdown* AND covid | 966 |
| #4 | First 10 pages | 100 |

PRISMA diagram for Tobacco smoking

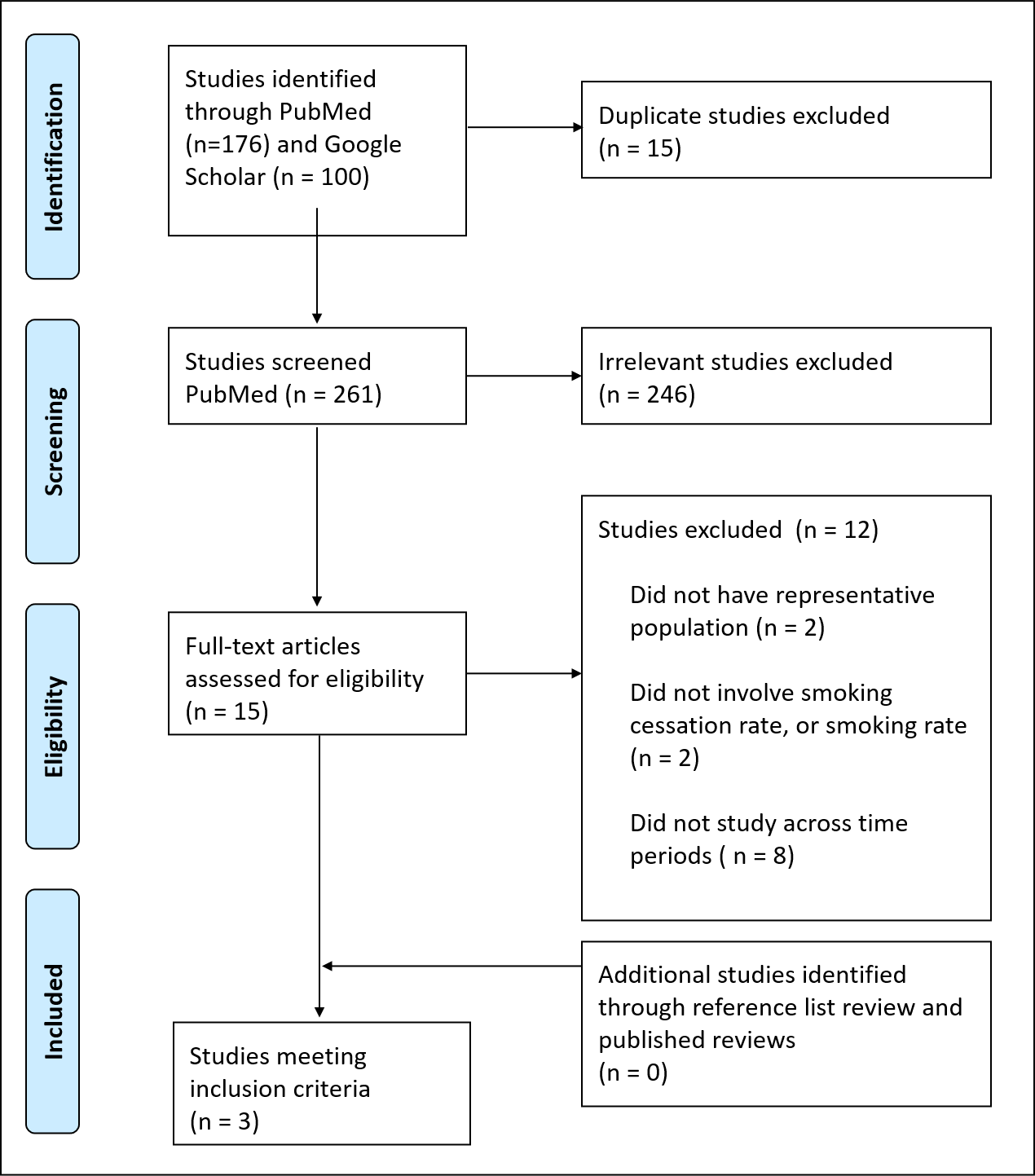

##### Alcohol consumption

**Literature search criteria (PICO framework)**

| **Inclusion criteria** | |
| --- | --- |
| Population | People in OECD countries, as well as China and HK, and Singapore. |
| Intervention | COVID 19 management related policies |
| Comparators | Different stages of the policy intervention |
| Outcome | Risks associated with changes in alcohol consumption patterns |
| Language restrictions | None |
| Search dates | Restricted to literature published between 1 January 2020 and 21 January 2021 |
| **Exclusion criteria** | |
| Population | Non-Human based studies… |
| Study design | Case Studies, surveys with no comparison… |

**Database Search Terms**

**Pubmed**

| **ID** | **Search** | **Hits** |
| --- | --- | --- |
| #1 | (((quarantine*  OR quarantine*  OR lockdown  OR lockdown  OR "policy response*"  OR "closure*"  OR "policy measure*" ) ) AND (alcohol) | 105 |
| #2 | ((quarantine*  OR quarantine*  OR lockdown  OR lockdown  OR "policy response*"  OR "closure*"  OR "policy measure*" ) AND (alcohol impact)) AND (australia) | 51 |
| #3 | **((quarantine*  OR quarantine*  OR lockdown  OR lockdown  OR "policy response*"  OR "closure*"  OR "policy measure*" ) AND (alcohol impact))** | **33** |
| #4 | ((quarantine*  OR quarantine*  OR lockdown  OR lockdown  OR "policy response*"  OR "closure*"  OR "policy measure*" ) AND (alcohol impact)) AND injury | 2 |
| #5 | (((quarantine*  OR quarantine*  OR lockdown  OR lockdown  OR "policy response*"  OR "closure*"  OR "policy measure*" ) AND (alcohol impact)) AND (alcohol ) | 37 |

**Google Scholar**

Search date 23 Jan 2021. Restricted to articles published since 1 January 2020 and sorted by relevance. The first ten pages of each search was assessed due to the large number of hits.

| **ID** | **Search** | **Hits** |
| --- | --- | --- |
| 1 | Coronavirus | 1, 240,000 |
| 2 | Quarantine OR lockdown OR isolation OR “policy responses” | 3,610,000 |
| 3 | 1 AND 2 | 263,400 |
| 4 | Alcohol AND 3 | 41 000 |
| 5 | Alcohol habits OR behaviours and 4 | 4510 |
| 6 | Alcoholism and 5 | 2004 |
| 7 | First 10 pages | 100 |

PRISMA Diagram for Alcohol

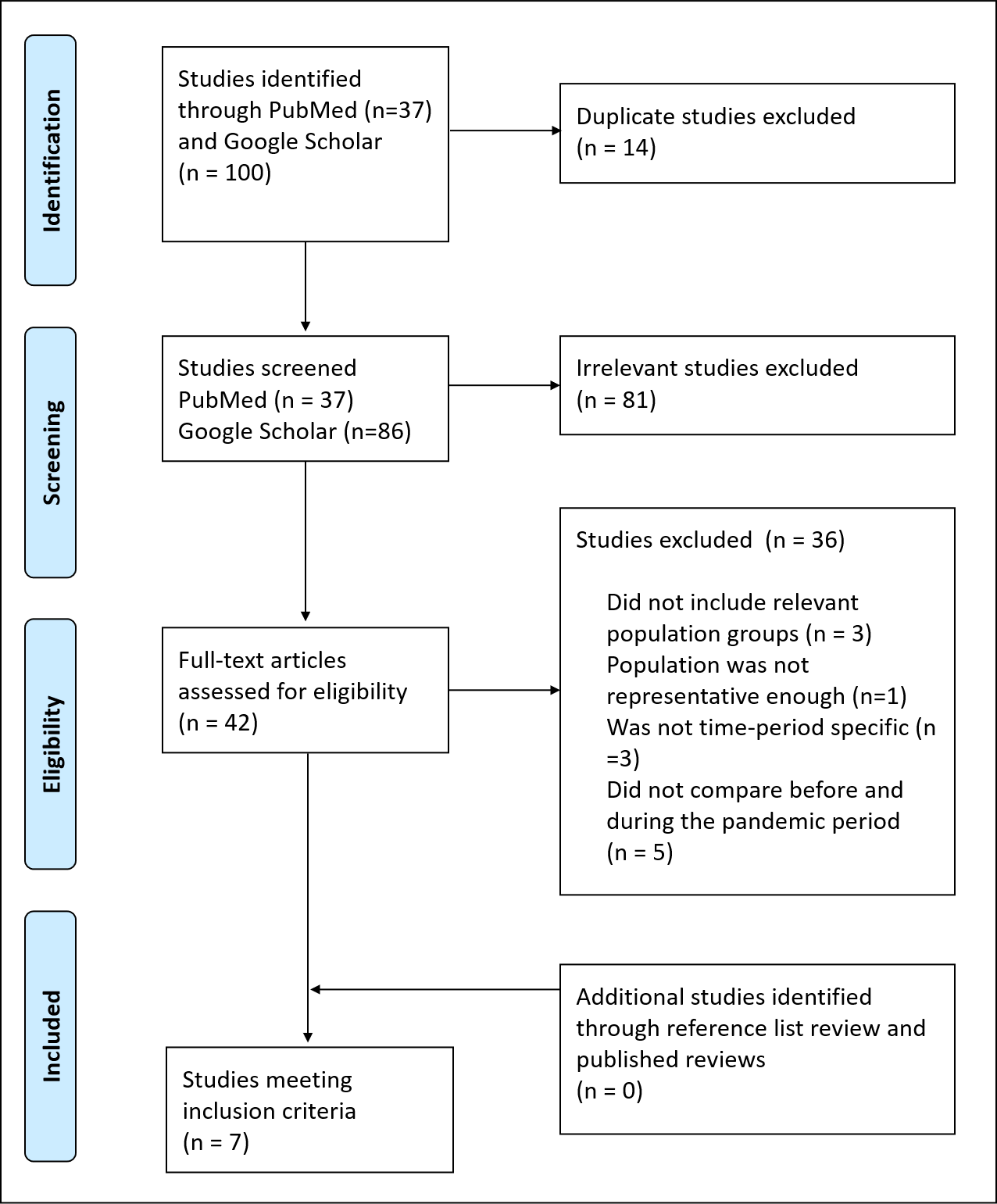

##### Physical activity (PA)

**Literature search criteria (PICO framework)**

| **Inclusion criteria** | |
| --- | --- |
| Population | Australia   Australia and OECD countries plus China and Singapore |
| Intervention | Several policy measures for COVID-19 pandemic |
| Comparators | Scenario without the policy measure |
| Outcome | Change in Minutes of Moderate to Vigorous Physical Activity per `time’ |
| Language restrictions | None |
| Search dates | Restricted to literature published between 1 January 2020 and 21 January 2021 |
| **Exclusion criteria** | |
| Population | Non-Human based studies… |
| Study design | Case Studies, surveys with no comparison…, No measure of physical activity |

**Database Search Terms**

**Pubmed**

| **ID** | **Search** | **Hits** |
| --- | --- | --- |
| 1 | coronavirus*[mh] OR coronavirus*[tw] OR coronavirus*[tiab] OR COVID-19[tw] OR COVID*[tiab] | 34,789 |
| 2 | quarantine*[tiab] OR quarantine*[tw] OR lockdown[tiab] OR lockdown[tw] OR "response*"[tiab] OR "closure*"[tiab] OR "policy measure*"[tiab] OR "confinement"[tiab] or home[tiab] OR social distancing[tiab] OR isolation[tiab] OR restrictions*[tiab] | 75,088 |
| 3 | 1 AND 2^1^ | 9,715 |
| 4 | Physical Activity and 3 | 447 |
| 6 | “physical activity patterns”OR rates AND 4 | 155 |

**Google Scholar**

Search date January 20, 2021. Restricted to articles published since 1 January 2020 and sorted by relevance. The first ten pages of each search was assessed due to the large number of hits.

| **ID** | **Search** | **Hits** |
| --- | --- | --- |
| 1 | Coronavirus | 1,240,000 |
| 2 | Quarantine OR lockdown OR isolation OR “policy responses” | 3,610,000 |
| 3 | 1 AND 2 | 273,400 |
| 4 | 3 AND Australia | 58 000 |
| 5 | Physical Activity and 4 | 23 000 |
| 6 | “Time series” AND 5 | 17 300 |
| 7 | Exercise and 6 | 7320 |
| 8 | First 10 pages | 100 |

 PRISMA diagram for Physical Activity

  
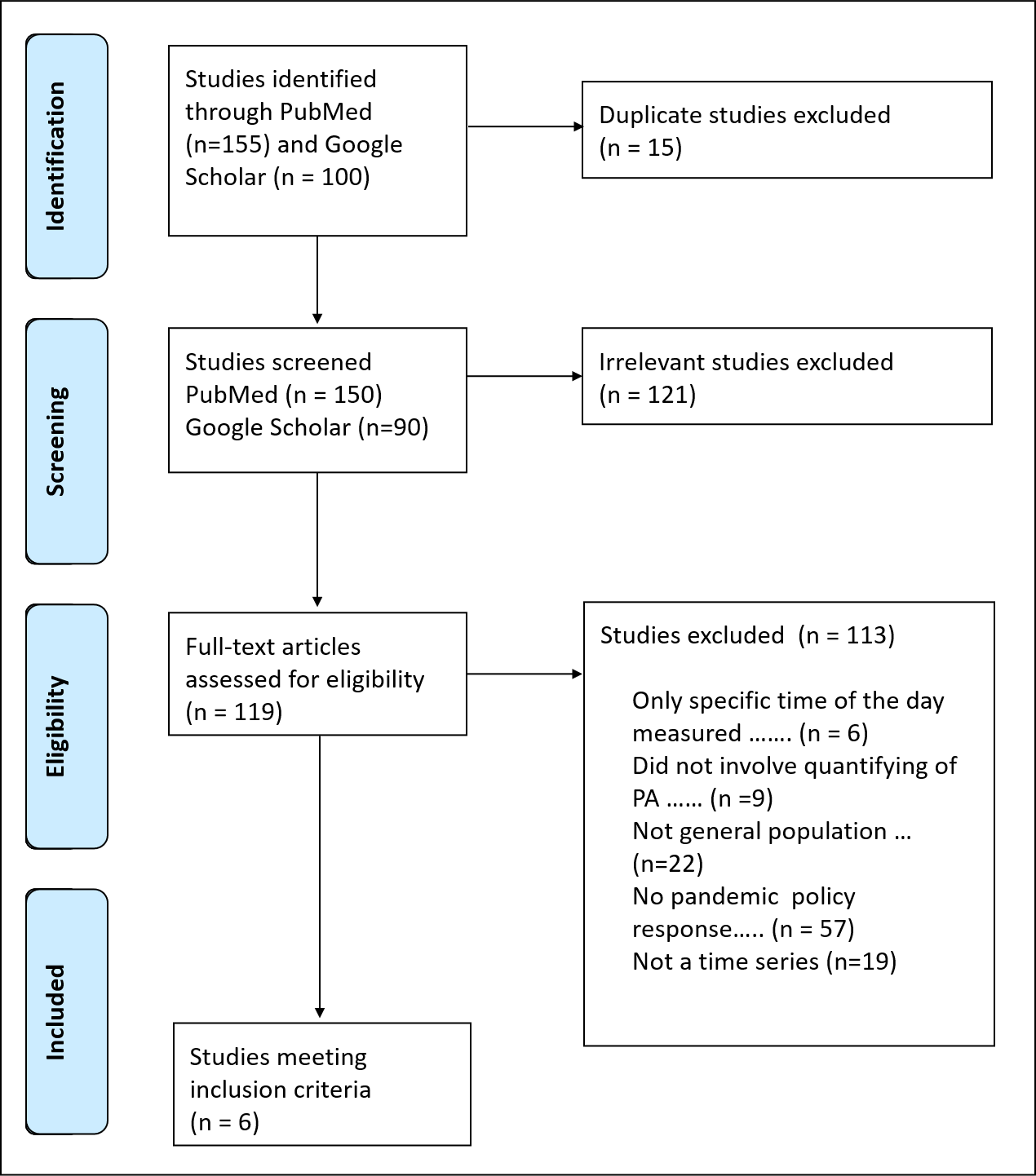

##### Intimate partner violence (IPV)

**Literature search criteria (PICO framework)**

| **Inclusion criteria** | |
| --- | --- |
| Population | People in OECD countries, as well as China and HK, and Singapore. |
| Intervention | COVID 19 management related policies |
| Comparators | Different stages of the policy intervention |
| Outcome | Changes in intimate partner violence |
| Language restrictions | None |
| Search dates | Restricted to literature published between 1 January 2020 and 21 January 2021 |
| **Exclusion criteria** | |
| Population | Non-Human based studies… |
| Study design | Case Studies, surveys with no comparison… |

**Database Search Terms**

**Pubmed**

| **ID** | **Search** | **Hits** |
| --- | --- | --- |
| #1 | intimate partner violence AND lockdown* | 11 |
| #2 | (intimate partner violence OR IPV) AND (lockdown* OR stay-at-home) | 19 |
| #3 | (intimate partner violence OR IPV) AND (Australia) AND (Covid OR Lockdown) | 4 |
| #4 | (Lockdown* OR Stay-at-Home*) AND (Intimate partner violence OR IPV OR Violence Against Women) | 25 |
| **#5** | **(Intimate partner violence OR IPV OR Violence Against Women) AND (Covid)** | 89 |
| #6 | (Intimate partner violence OR IPV OR Violence Against Women OR Domestic Violence) AND (Covid) | 239 |

**Google Scholar**

Search date 22^nd^ January, 2021. Restricted to articles published since 1 January 2020 and sorted by relevance. The first ten pages of each search was assessed due to the large number of hits.

| **ID** | **Search** | **Hits** |
| --- | --- | --- |
| #1 | intimate partner violence AND lockdowns | 2280 |
| #2 | intimate partner violence AND lockdowns AND Australia | 884 |
| #3 | (intimate partner violence OR domestic violence) AND Covid AND Australia | 2560 |
| #4 | First 10 pages | 100 |

PRISMA diagram for IPV

  
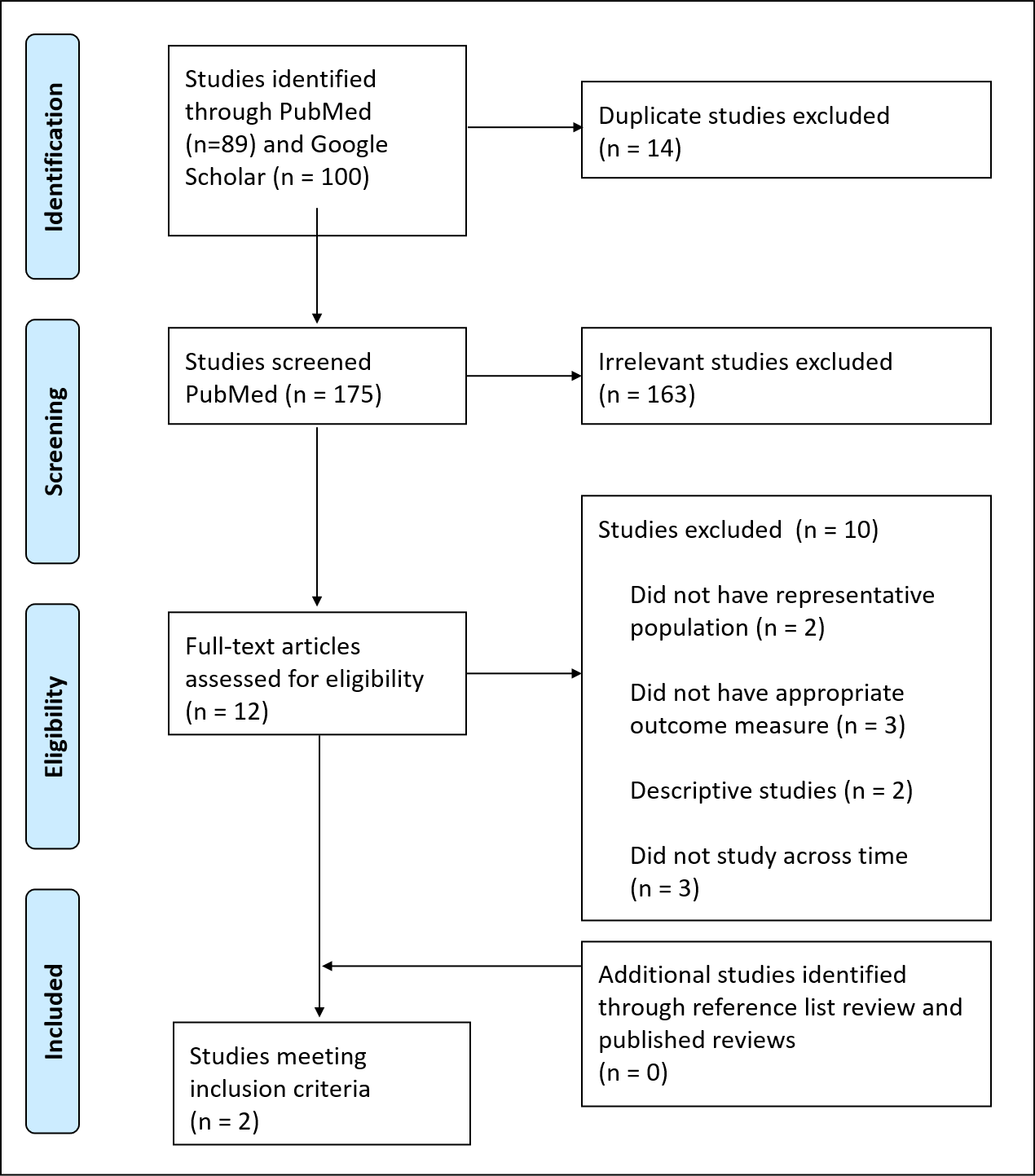

##### Suicide

**Literature search criteria (PICO framework)**

| **Inclusion criteria** | |
| --- | --- |
| Population | Australia   Australia and OECD countries plus China and Singapore |
| Intervention | Several policy measures for COVID-19 pandemic |
| Comparators | Scenario without the policy measure |
| Outcome | Change in Suicide trends |
| Language restrictions | None |
| Search dates | Restricted to literature published between 1 January 2020 and 22 January 2021 |
| **Exclusion criteria** | |
| Population | Non-Human based studies… |
| Study design | Case Studies, time series data comparisons |

**Database Search Terms**

**Pubmed**

| **ID** | **Search** | **Hits** |
| --- | --- | --- |
| 1 | coronavirus*[mh] OR coronavirus*[tw] OR coronavirus*[tiab] OR COVID-19[tw] OR COVID*[tiab] | 34,789 |
| 2 | quarantine*[tiab] OR quarantine*[tw] OR lockdown[tiab] OR lockdown[tw] OR "response*"[tiab] OR "closure*"[tiab] OR "policy measure*"[tiab] OR "confinement"[tiab] or home[tiab] OR social distancing[tiab] OR isolation[tiab] OR restrictions*[tiab] | 75,088 |
| 3 | 1 AND 2^1^ | 9,715 |
| 4 | Suicide AND 3 | 189 |
| 6 | Trends AND 4 | 25 |

**Google Scholar**

Search date 23^rd^ January 2021. Restricted to articles published since 1 January 2020 and sorted by relevance. The first ten pages of each search was assessed due to the large number of hits.

| **ID** | **Search** | **Hits** |
| --- | --- | --- |
| 1 | Coronavirus | 1, 240,000 |
| 2 | Quarantine OR lockdown OR isolation OR “policy responses” | 3,610,000 |
| 3 | 1 AND 2 | 273,400 |
| 4 | 3 AND Australia | 58 000 |
| 5 | Suicide and 4 | 2000 |
| 6 | “Time series” AND 5 | 1630 |
| 8 | First 10 pages | 100 |

 PRISMA diagram for suicide

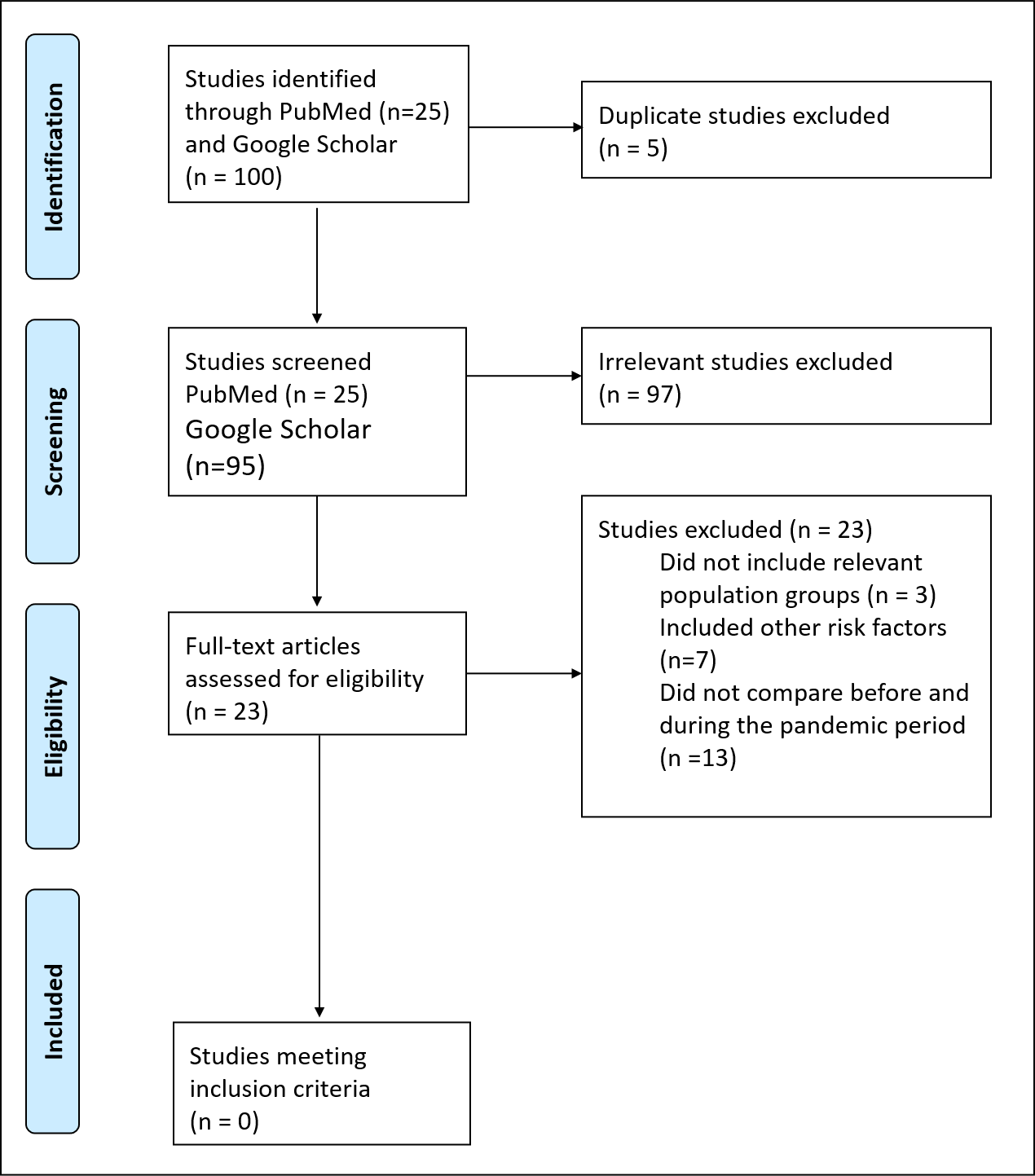

##### Body mass index (BMI)

**Literature search criteria (PICO framework)**

| **Inclusion criteria** | |
| --- | --- |
| Population | Australia   Australia and OECD countries plus China and Singapore |
| Intervention | Several policy measures for COVID-19 pandemic |
| Comparators | Scenario without the policy measure |
| Outcome | Change in BMI |
| Language restrictions | None |
| Search dates | Restricted to literature published between 1 January 2020 and 22 January 2021 |
| **Exclusion criteria** | |
| Population | Non-Human based studies… |
| Study design | Case Studies, surveys with no comparison… |

**Database Search Terms**

**Pubmed**

| **ID** | **Search** | **Hits** |
| --- | --- | --- |
| 1 | coronavirus*[mh] OR coronavirus*[tw] OR coronavirus*[tiab] OR COVID-19[tw] OR COVID*[tiab] | 34,789 |
| 2 | quarantine*[tiab] OR quarantine*[tw] OR lockdown[tiab] OR lockdown[tw] OR "response*"[tiab] OR "closure*"[tiab] OR "policy measure*"[tiab] OR "confinement"[tiab] or home[tiab] OR social distancing[tiab] OR isolation[tiab] OR restrictions*[tiab] | 75,088 |
| 3 | 1 AND 2^1^ | 9,715 |
| 4 | “body mass index” AND 3 | 51 |
| 5 | BMI AND 4 | 25 |

**Google Scholar**

Search date 22^nd^ January, 2021. Restricted to articles published since 1 January 2020 and sorted by relevance. The first ten pages of each search was assessed due to the large number of hits.

| **ID** | **Search** | **Hits** |
| --- | --- | --- |
| 1 | Coronavirus | 1, 240,000 |
| 2 | Quarantine OR lockdown OR isolation OR “policy responses” | 3,610,000 |
| 3 | 1 AND 2 | 263,400 |
| 4 | BMI and 3 | 21 000 |
| 5 | “Time series” AND 4 | 16 000 |
| 6 | Body Mass Index AND 5 | 12 000 |
| 7 | Lockdown impact AND 6 | 2210 |
| 8 | First 10 pages | 100 |

  PRISMA diagram for BMI

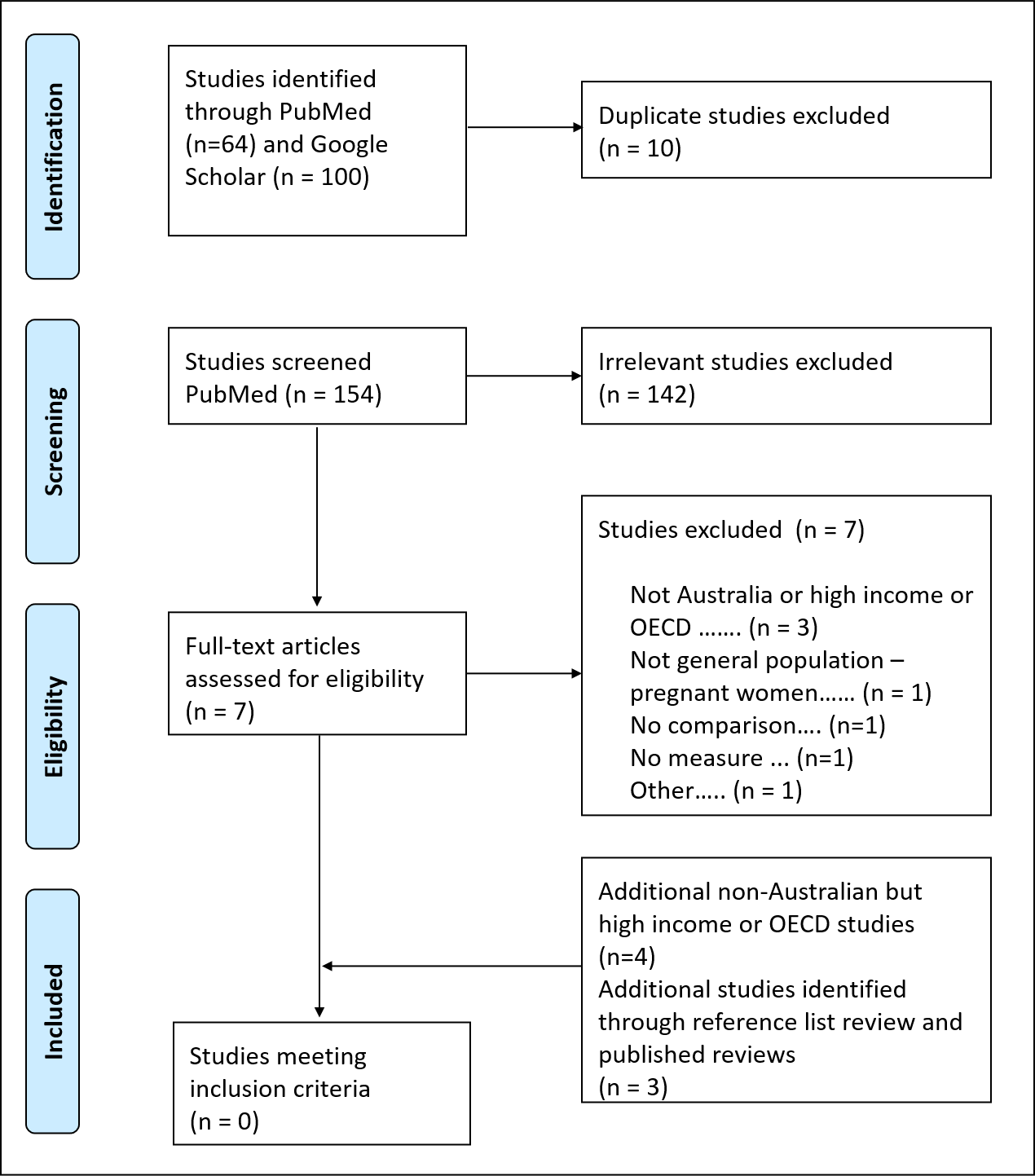

##### Stroke

**Literature search criteria (PICO framework)**

| **Inclusion criteria** | |
| --- | --- |
| Population | Studies conducted in OECD countries, China, Honk Kong, Singapore. |
| Intervention | Several policy measures for COVID-19 pandemic |
| Comparators | Scenario without the policy measure |
| Outcome | Incidence of Stroke |
| Language restrictions | None |
| Search dates | Restricted to literature published between 1 January 2020 and 22 January 2021 |
| **Exclusion criteria** | |
| Population | Non-Human based studies… |
| Study design | Case Studies, surveys with no comparison… |

**Database Search Terms**

**Pubmed**

| **ID** | **Search** | **Hits** |
| --- | --- | --- |
| 1 | coronavirus*[mh] OR coronavirus*[tw] OR coronavirus*[tiab] OR COVID-19[tw] OR COVID*[tiab] | 119 000 |
| 2 | quarantine*[tiab] OR quarantine*[tw] OR lockdown[tiab] OR lockdown[tw] OR “response*”[tiab] OR “closure*”[tiab] OR “policy measure*”[tiab]  OR “confinement”[tiab] or home[tiab] OR distancing[tiab]  OR isolation[tiab]  OR restrictions*[tiab] | 3, 095,048 |
| 3 | 1 AND 2 | 29,358 |
| 4 | Stroke | 362,179 |
| 5 | 3 AND 4 | 320 |
| 6 | 5 AND lockdown | 29 |

**Google Scholar**

Search date 22^nd^ January, 2021. Restricted to articles published since 1 January 2020 and sorted by relevance. The first ten pages of each search was assessed due to the large number of hits.

| **ID** | **Search** | **Hits** |
| --- | --- | --- |
| 1 | Coronavirus | 158,000 |
| 2 | Quarantine OR lockdown OR isolation OR “policy responses” | 110,000 |
| 3 | 1 AND 2^6^ | 58,400 |
| 4 | Australia | - |
| 5 | 3 AND 4^7^ | 3,910 |
| 6 | stroke | 54,000 |
| 7 | 5 AND 6^8^ | 1,060 |
| 6 | First 10 pages | 100 |

  PRISMA diagram for Stroke

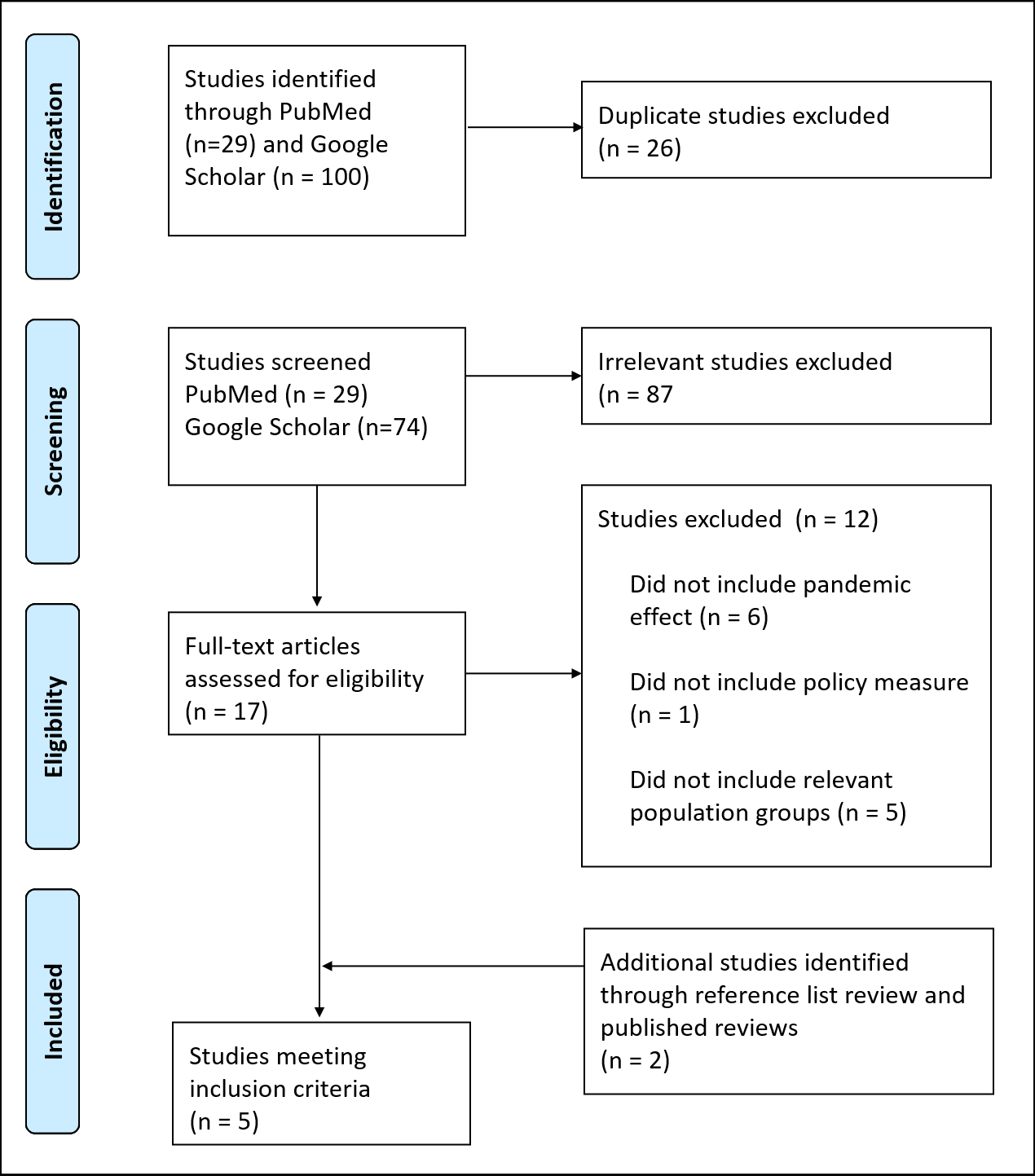

##### Anxiety disorders

**Literature search criteria (PICO framework)**

| **Inclusion criteria** | |
| --- | --- |
| Population | Studies conducted in OECD countries, China, Honk Kong, Singapore. |
| Intervention | COVID 19 management related policies |
| Comparators | Different stages of the policy intervention |
| Outcome | Changes in tobacco consumption |
| Language restrictions | None |
| Search dates | Restricted to literature published between 1 January 2020 and 22 January 2021 |
| **Exclusion criteria** | |
| Population | Non-Human based studies… |
| Study design | Case Studies, surveys with no comparison… |

**Database Search Terms**

**Pubmed**

| **ID** | **Search** | **Hits** |
| --- | --- | --- |
| #1 | (anxiety OR anxiety disorders) | 25088 |
| #2 | #1 AND Australia* | 1767 |
| #3 | #2 AND (lockdown* OR quarantine OR Covid policy) | 47 |
| #4 | **anxiety OR anxiety disorders AND Australia* AND (lockdown* OR quarantine OR Covid policy)** | 47 |

**Google Scholar**

Search date 22^nd^ January, 2021. Restricted to articles published since 1 January 2020 and sorted by relevance. The first ten pages of each search was assessed due to the large number of hits.

| **ID** | **Search** | **Hits** |
| --- | --- | --- |
| #1 | lockdown* OR stay-at-home OR stay at home AND anxiety OR anxiety disorder* AND Australia | 4020 |
| #2 | anxiety disorder AND Australia lockdown | 2610 |
| #3 | **anxiety AND Australia lockdown AND covid** | 4790 |
| #4 | anxiety levels AND Australia lockdown | 4550 |
| #5 | First 10 pages | 100 |

 PRISMA diagram for Anxiety disorders

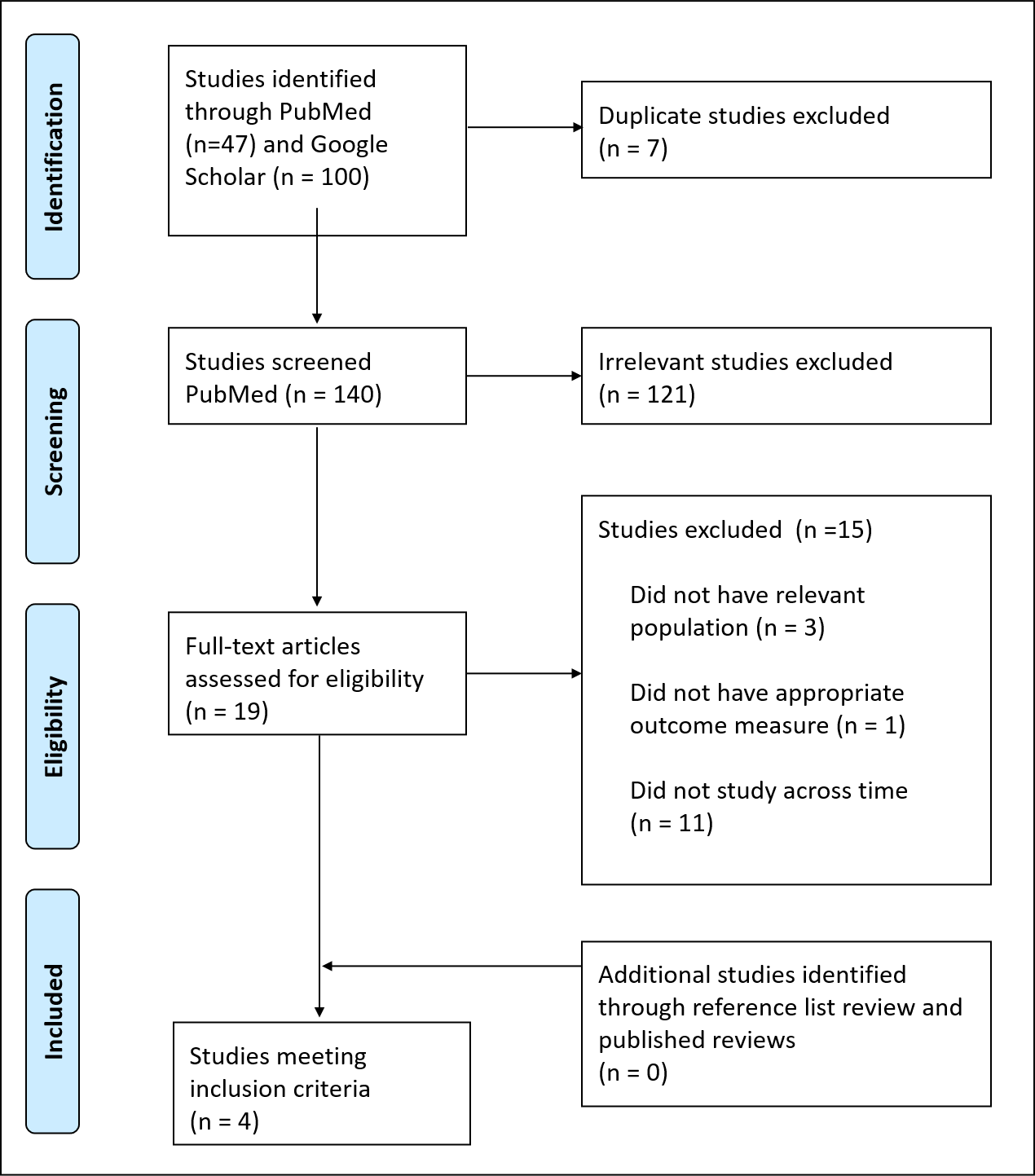

##### Depression

Literature search criteria (PICO framework)

| **Inclusion criteria** | |
| --- | --- |
| Population | Studies conducted in OECD countries, China, Honk Kong, Singapore. |
| Intervention | COVID 19 management related policies |
| Comparators | Different stages of the policy intervention |
| Outcome | Changes in tobacco consumption |
| Language restrictions | None |
| Search dates | Restricted to literature published between 1 January 2020 and 22 January 2021 |
| **Exclusion criteria** | |
| Population | Non-Human based studies… |
| Study design | Case Studies, surveys with no comparison… |

**Database Search Terms**

**Pubmed**

| **ID** | **Search** | **Hits** |
| --- | --- | --- |
| #1 | (depression OR depressive disorders OR dysthymia) | 37727 |
| #2 | #1 AND Australia | 2380 |
| #3 | #2 AND (**lockdown* OR quarantine OR Covid policy)** | **41** |
| #4 | (lockdown* OR Covid policy) AND (depression OR depressive disorders OR dysthymia) AND Australia | 38 |
| #5 | lockdown* OR quarantine OR Covid policy) AND (depression OR depressive disorders OR dysthymia) | 772 |

**Google Scholar**

Search date 22^nd^ January, 2021. Restricted to articles published since 1 January 2020 and sorted by relevance. The first ten pages of each search was assessed due to the large number of hits.

| **ID** | **Search** | **Hits** |
| --- | --- | --- |
| #1 | lockdown* OR stay-at-home OR stay at home AND depression OR major depressive disorder* AND Australia | 1100 |
| #2 | major depressive disorder AND Australia lockdown | 1710 |
| #3 | major depressive disorder AND Australia lockdown AND covid | 3210 |
| #4 | First 10 pages | 100 |

 PRISMA diagram for Depression

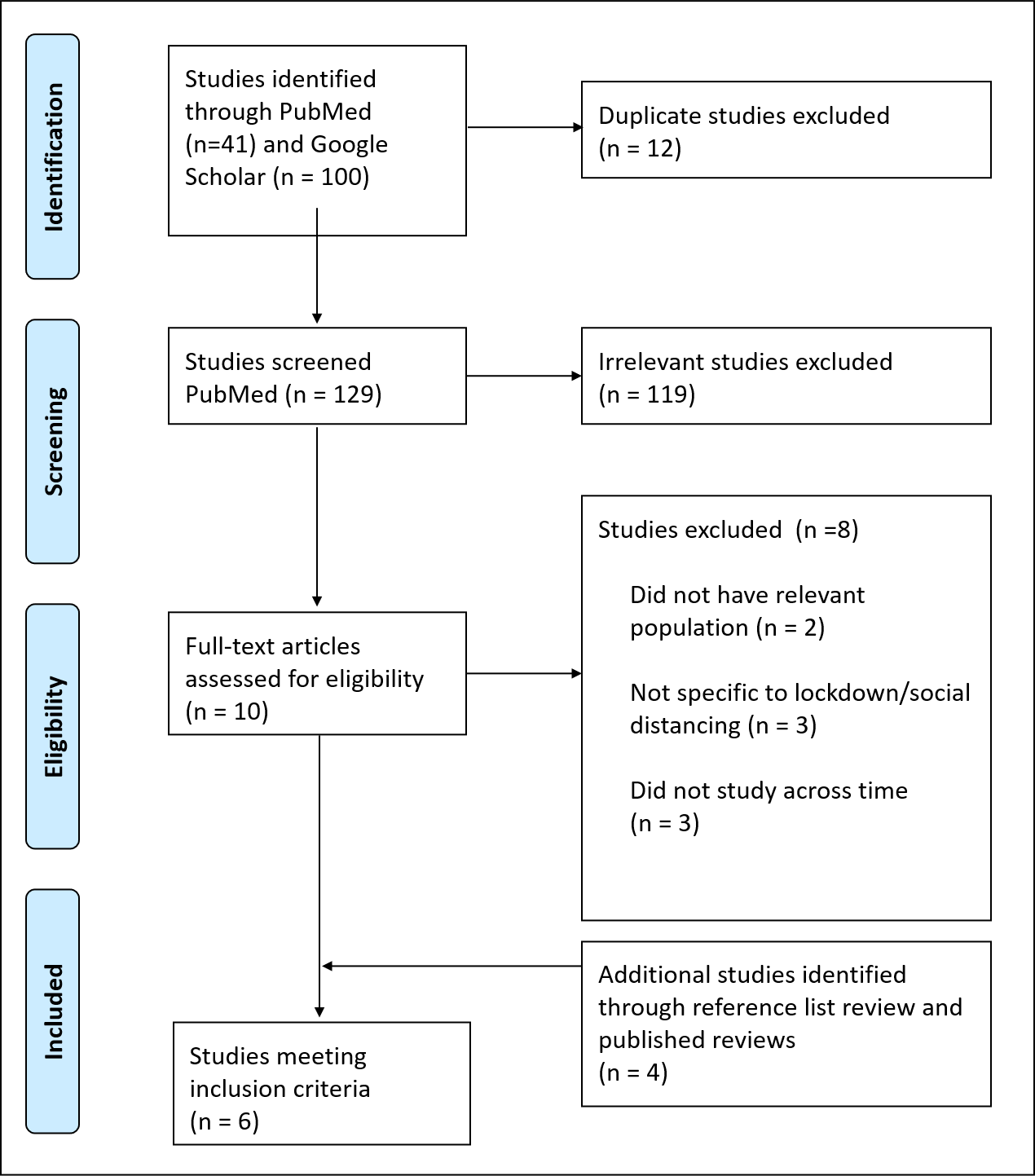

##### Road transport injuries (RTI)

Literature search criteria (PICO framework)

| **Inclusion criteria** | |
| --- | --- |
| Population | Studies conducted in OECD countries, China, Honk Kong, Singapore. |
| Intervention | COVID 19 management related policies |
| Comparators | Different stages of the policy intervention |
| Outcome | Change in RTI indidence during the lockdown |
| Language restrictions | English |
| Search dates | Restricted to literature published between 1 January 2020 and 22 January 2021 |
| **Exclusion criteria** | |
| Population | Non-Human based studies… |
| Study design | Case Studies, surveys with no comparison… |

**Database Search Terms**

**Pubmed**

| **ID** | **Search** | **Hits** |
| --- | --- | --- |
| #1 | (road accident) OR (vehicle accident) OR (car accident) AND (COVID19) | 32 |
| #2 | (road accident) OR (vehicle accident) OR (car accident) AND (lockdown) | 8 |
| #3 | (road accident) OR (vehicle accident) OR (car accident) AND (COVID19) AND (policy) | 6 |

**Google Scholar**

Search date 22^nd^ January, 2021. Restricted to articles published since 1 January 2020 and sorted by relevance. The first ten pages of each search was assessed due to the large number of hits.

| **ID** | **Search** | **Hits** |
| --- | --- | --- |
| #1 | Road (OR) vehicle accidents (and) covid19 policy | 19300 |
| #2 | car accidents (OR) road traffic accidents (and) covid19 lockdown | 3230 |
| #3 | road traffic accidents (and) covid 19 (and) Australia | 974 |
| #4 | road traffic accidents (and) covid 19 policy (and) Australia | 880 |
| #5 | road traffic accidents (and) covid19 (and) lockdown | 743 |
| #6 | vehicle accidents (and) covid 19 (and) lockdown (and) Australia | 629 |
| #7 | road traffic accidents (and) covid19 lockdown policy (and) Australia | 270 |
| #8 | First 10 pages | 100 |

PRISMA diagram for Road Transport Injuries

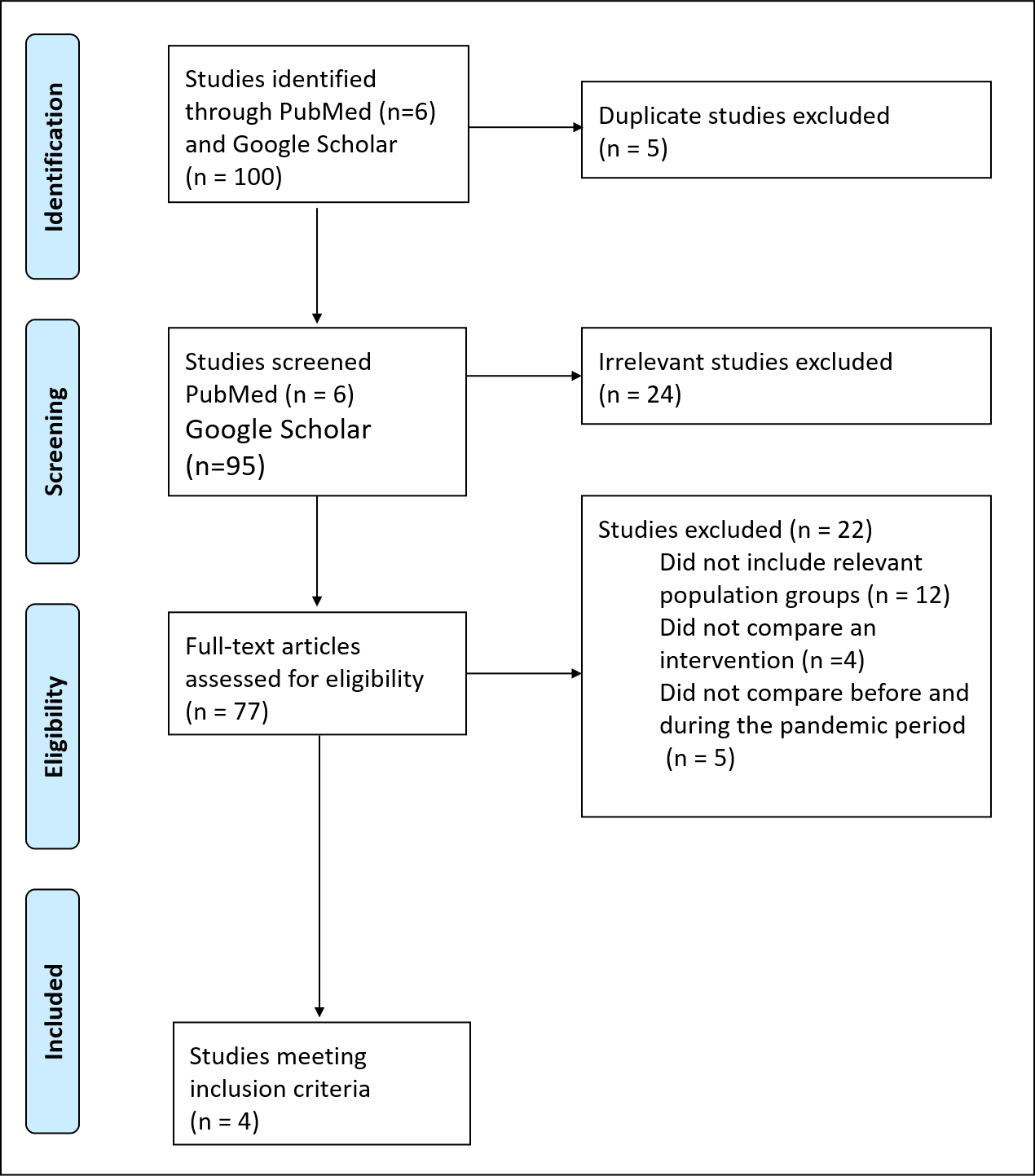

##### Falls

Literature search criteria (PICO framework)

| **Inclusion criteria** | |
| --- | --- |
| Population | Studies conducted in OECD countries, China, Honk Kong, Singapore. |
| Intervention | COVID 19 management related policies |
| Comparators | Different stages of the policy intervention |
| Outcome | Falls or injuries amongst the elderly |
| Language restrictions | None |
| Search dates | Restricted to literature published between 1 January 2020 and 22 January 2021 |
| **Exclusion criteria** | |
| Population | Non-Human based studies… |
| Study design | Case Studies, surveys with no comparison… |

**Database Search Terms**

**Pubmed**

| **ID** | **Search** | **Hits** |
| --- | --- | --- |
| 1 | falls | 94093 |
| 3 | falls and injuries | 22291 |
| 3 | (falls and injuries) AND (covid19) AND (SARS-CoV-2) | 13 |

**Google Scholar**

Search date 22^nd^ January, 2021. Restricted to articles published since 1 January 2020 and sorted by relevance. The first ten pages of each search was assessed due to the large number of hits.

| **ID** | **Search** | **Hits** |
| --- | --- | --- |
| #1 | falls and instability in the elderly | 94100 |
| #2 | falls and injuries 0R instability in the elderly | 35700 |
| #3 | falls and injuries or instability in the elderly AND covid19 | 7730 |
| #4 | falls and injuries or instability in the elderly AND covid19 AND lockdown | 1330 |
| #5 | falls and injuries or instability in the elderly AND covid19 (AND) lockdown (OR) COVID19 policy | 1210 |
| #6 | falls and injuries or instability in the elderly AND covid19 (AND) lockdown (OR) COVID19 policy (AND) Australia | 629 |
| #7 | First 10 pages only | 100 |

 PRISMA diagram for Falls

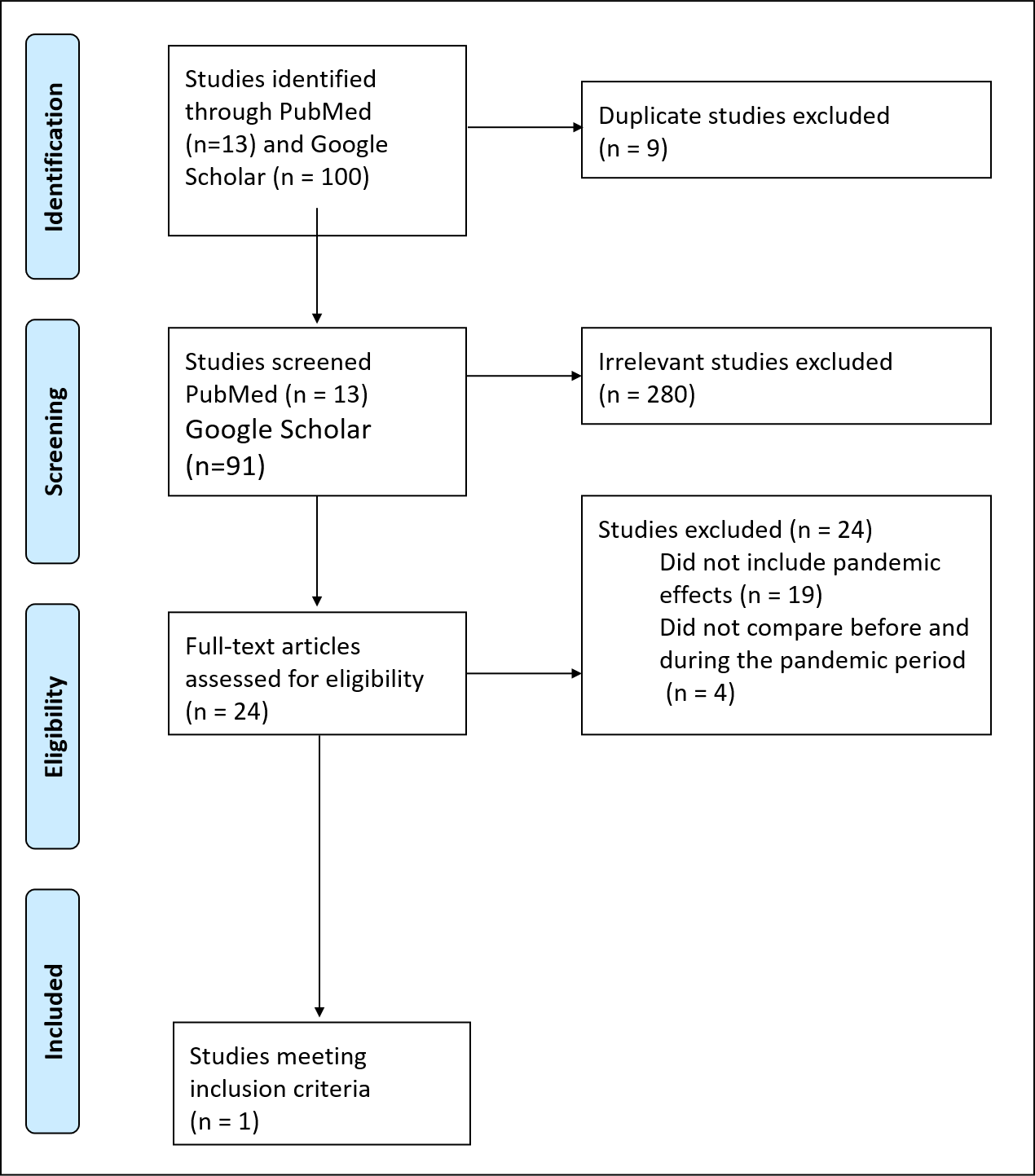

##### Self-harm/Interpersonal violence (IV)

Literature search criteria (PICO framework)

| **Inclusion criteria** | |
| --- | --- |
| Population | Studies conducted in OECD countries, China, Honk Kong, Singapore. |
| Intervention | COVID 19 management related policies |
| Comparators | Different stages of the policy intervention |
| Outcome |  |
| Language restrictions | None |
| Search dates | Restricted to literature published between 1 January 2020 and 22 January 2021 |
| **Exclusion criteria** | |
| Population | Non-Human based studies… |
| Study design | Case Studies, surveys with no comparison… |

**Database Search Terms**

**Pubmed**

| **ID** | **Search** | **Hits** |
| --- | --- | --- |
| #1 | **(self harm OR interpersonal violence) AND (lockdown* OR stay at home OR quarantine) AND (covid)** | **84** |
| #2 | (self harm OR interpersonal violence) AND (lockdown* OR stay at home OR quarantine) AND (Australia) | 6 |
| #3 | (self harm) AND (lockdown* OR stay at home OR stay-at-home) | 50 |
| #4 | self harm AND Covid | 261 |
| #5 | **interpersonal violence AND Covid** | **28** |
| #6 | interpersonal violence AND lockdown | 7 |

**Google Scholar**

Search date 22^nd^ January, 2021. Restricted to articles published since 1 January 2020 and sorted by relevance. The first ten pages of each search was assessed due to the large number of hits.

| **ID** | **Search** | **Hits** |
| --- | --- | --- |
| #1 | **self harm OR interpersonal violence AND lockdown* OR stay at home ANDAustralia** | **4110** |
| #2 | self harm AND covid lockdown* AND Australia | 4860 |
| #3 | interpersonal violence AND covid lockdown* AND Australia | 959 |
| #4 | First 10 pages | 100 |

PRISMA diagram for Self-harm/Interpersonal violence (IV)

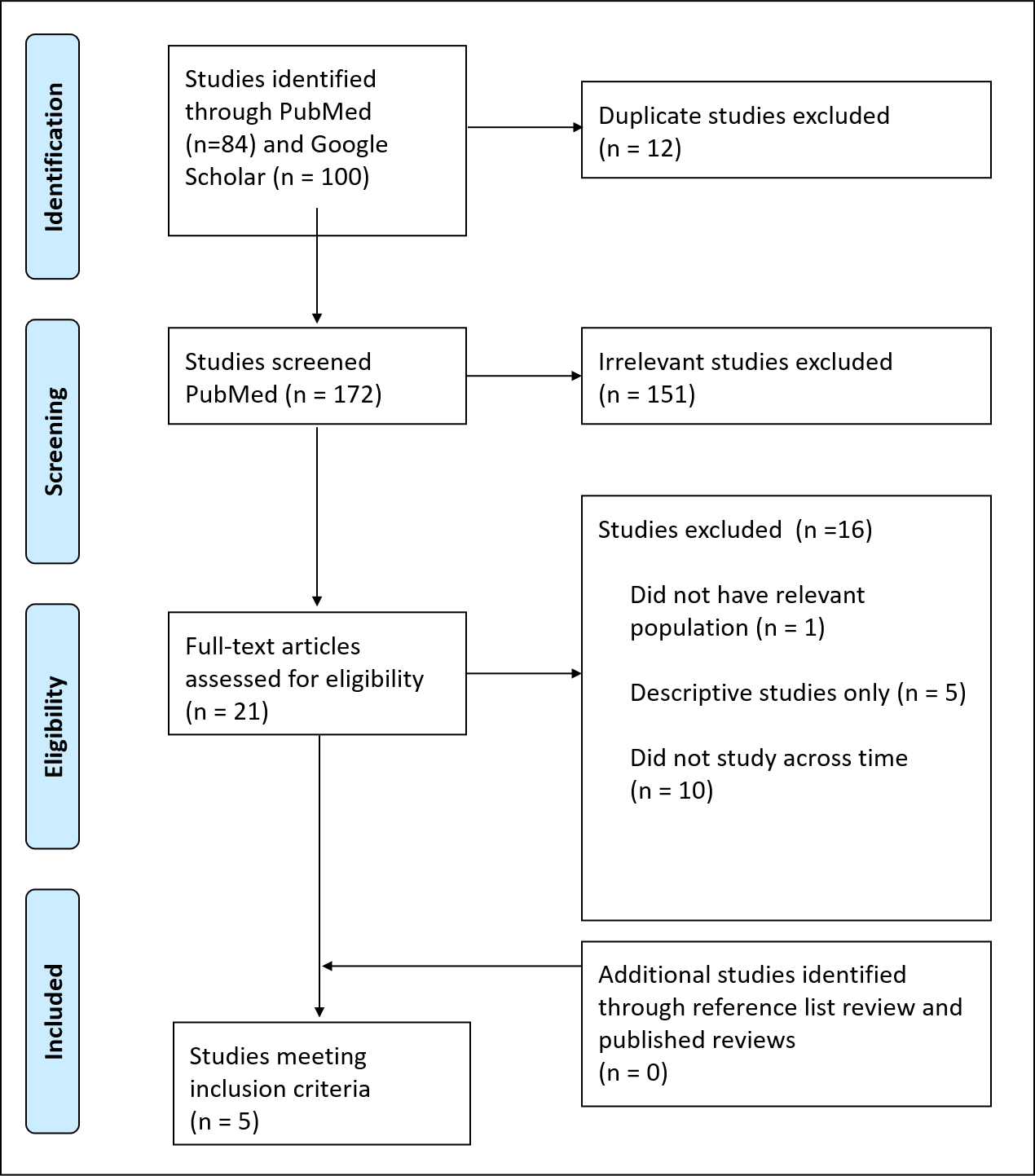

##### Cancer screening

Literature search criteria (PICO framework)

| **Inclusion criteria** | |
| --- | --- |
| Population | Studies conducted in OECD countries, China, Honk Kong, Singapore. |
| Intervention | COVID 19 management related policies |
| Comparators | Different stages of the policy intervention |
| Outcome | Screening participation for melanoma, breast, cervical, colorectal and lung cancer |
| Language restrictions | None |
| Search dates | Restricted to literature published between 1 January 2020 and 22 January 2021 |
| **Exclusion criteria** | |
| Population | Non-Human based studies… |
| Study design | Case Studies, cross-sectional surveys with no comparison |

**Database Search Terms**

**Pubmed**

| **ID** | **Search** | **Hits** |
| --- | --- | --- |
| #1 | (((Cancer screening) AND (Melanoma OR Breast OR Cervical OR Colorectal OR Lung)) AND (COVID-19 OR SARS-COV-2)) AND (Lockdown* OR "stay at home" OR "social distancing" OR "physical distancing" OR restrictions) | 47 |

**Google Scholar**

Search date 22^nd^ January, 2021. Restricted to articles published since 1 January 2020 and sorted by relevance. The first ten pages of each search was assessed due to the large number of hits.

| **ID** | **Search** | **Hits** |
| --- | --- | --- |
| #1 | (((Cancer screening) AND (Melanoma OR Breast OR Cervical OR Colorectal OR Lung)) AND (COVID-19 OR SARS-COV-2)) AND (Lockdown* OR "stay at home" OR "social distancing" OR "physical distancing" OR restrictions) | 5450 |

PRISMA diagram cancer screening

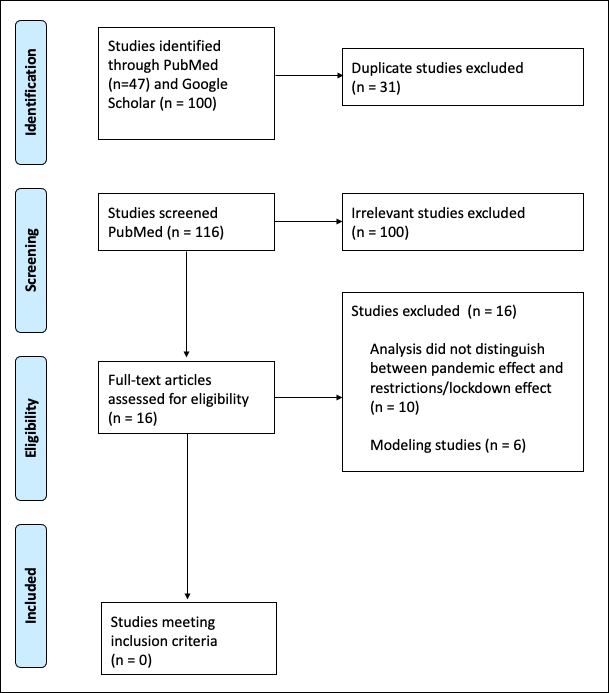

##### Ischaemic Heart Disease (IHD)

Literature search criteria (PICO framework)

| **Inclusion criteria** | |
| --- | --- |
| Population | Studies conducted in OECD countries, China, Honk Kong, Singapore. |
| Intervention | COVID 19 management related policies |
| Comparators | Different stages of the policy intervention |
| Outcome | Impact of pandemic response on IHD |
| Language restrictions | None |
| Search dates | Restricted to literature published between 1 January 2020 and 22 January 2021 |
| **Exclusion criteria** | |
| Population | Non-Human based studies… |
| Study design | Case Studies, surveys with no comparison… |

**Database Search Terms**

**Pubmed**

| **ID** | **Search** | **Hits** |
| --- | --- | --- |
| #1 | **ischaemic heart disease AND lockdown*** | 45 |
| #2 | (ischaemic heart disease OR heart failure OR IHD) AND lockdown* AND COVID-19 | 69 |
| #3 | (ischaemic heart disease OR heart failure OR IHD) AND COVID-19 | 1774 |
| #4 | **(ischaemic heart disease OR heart failure OR IHD) AND (lockdown* OR stay at home OR stay-at-home)** | **165** |
| #5 | (ischaemic heart disease OR heart failure OR IHD) AND (lockdown* OR stay at home OR stay-at-home) AND Australia | 5 |

**Google Scholar**

Search date 22^nd^ January, 2021. Restricted to articles published since 1 January 2020 and sorted by relevance. The first ten pages of each search was assessed due to the large number of hits.

| **ID** | **Search** | **Hits** |
| --- | --- | --- |
| #1 | ischaemic heart disease OR heart disease AND lockdown* OR stay at home AND Australia | **706** |
| #2 | ischaemic heart disease AND lockdown* OR stay at home | 2700 |
| #3 | ischaemic heart disease AND lockdown* OR stay at home AND Australia | 626 |
| #4 | First 10 pages | 100 |

PRISMA Diagram for ischaemic heart disease

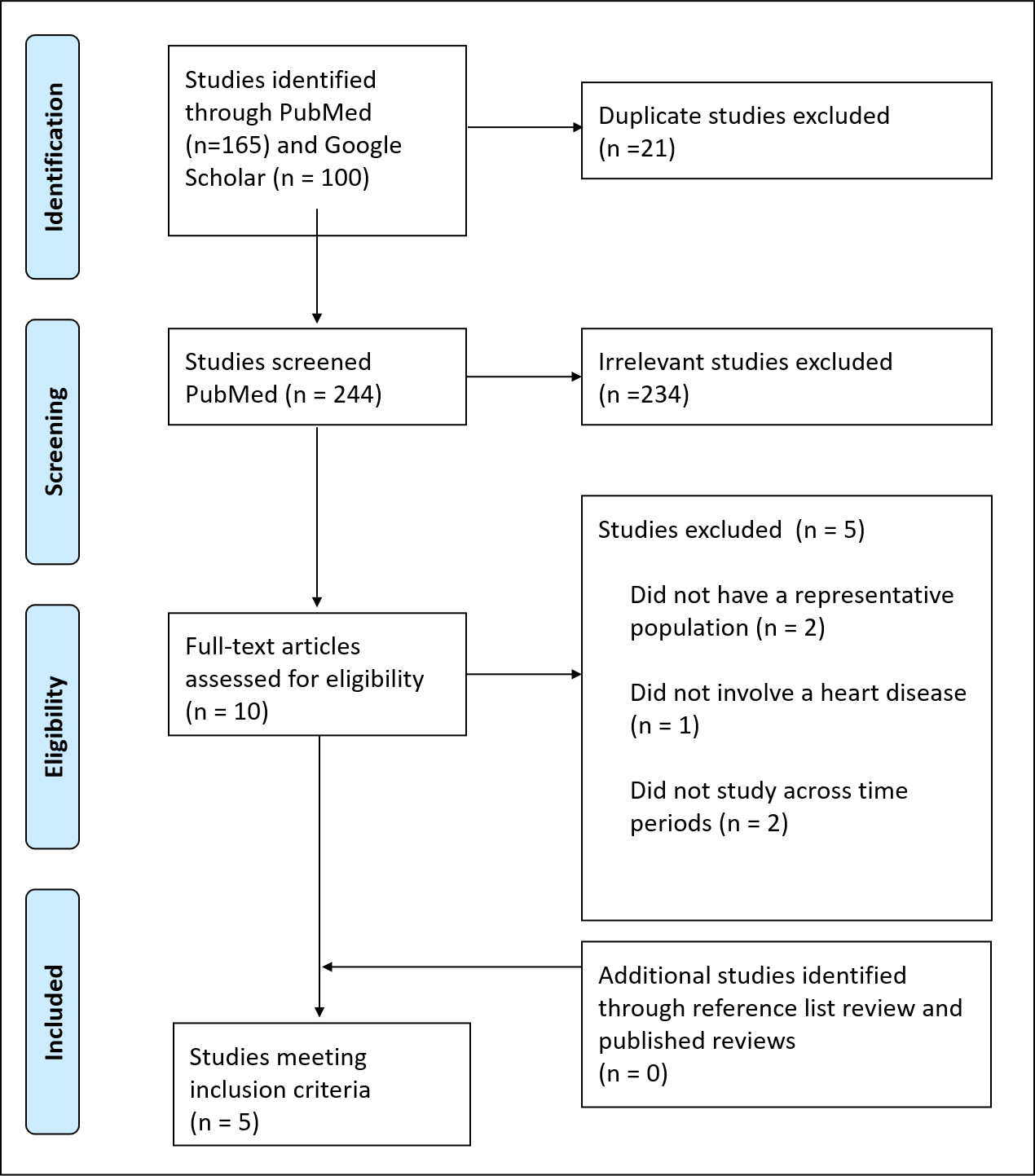

### **References**

1.	Munn Z, Peters MDJ, Stern C, Tufanaru C, McArthur A, Aromataris E. Systematic review or scoping review? Guidance for authors when choosing between a systematic or scoping review approach. BMC Medical Research Methodology. 2018;18(1):143.

2.	Niedzwiedz CL, Green MJ, Benzeval M, Campbell D, Craig P, Demou E, et al. Mental health and health behaviours before and during the initial phase of the COVID-19 lockdown: longitudinal analyses of the UK Household Longitudinal Study. J Epidemiol Community Health. 2020.

3.	Colbert S, Wilkinson C, Thornton L, Richmond R. COVID-19 and alcohol in Australia: Industry changes and public health impacts. Drug Alcohol Rev. 2020;39(5):435-40.

4.	Tran TD, Hammarberg K, Kirkman M, Nguyen HTM, Fisher J. Alcohol use and mental health status during the first months of COVID-19 pandemic in Australia. J Affect Disord. 2020;277:810-3.

5.	Rolland B, Haesebaert F, Zante E, Benyamina A, Haesebaert J, Franck N. Global Changes and Factors of Increase in Caloric/Salty Food Intake, Screen Use, and Substance Use During the Early COVID-19 Containment Phase in the General Population in France: Survey Study. JMIR Public Health Surveill. 2020;6(3):e19630.

6.	Dumas TM, Ellis W, Litt DM. What Does Adolescent Substance Use Look Like During the COVID-19 Pandemic? Examining Changes in Frequency, Social Contexts, and Pandemic-Related Predictors. Journal of Adolescent Health. 2020;67(3):354-61.

7.	Pellegrini M, Ponzo V, Rosato R, Scumaci E, Goitre I, Benso A, et al. Changes in Weight and Nutritional Habits in Adults with Obesity during the "Lockdown" Period Caused by the COVID-19 Virus Emergency. Nutrients. 2020;12(7).

8.	He M, Xian Y, Lv X, He J, Ren Y. Changes in Body Weight, Physical Activity, and Lifestyle During the Semi-lockdown Period After the Outbreak of COVID-19 in China: An Online Survey. Disaster Med Public Health Prep. 2020:1-6.

9.	Beigelman M, Vall Castelló J. COVID-19 and help-seeking behavior for intimate partner violence victims. IEB Working Paper 2020/13. 2020.

10.	Lundin R, Armocida B, Sdao P, Pisanu S, Mariani I, Veltri A, et al. Gender-based violence during the COVID-19 pandemic response in Italy. Journal of global health. 2020;10(2).

11.	Gallo LA, Gallo TF, Young SL, Moritz KM, Akison LK. The Impact of Isolation Measures Due to COVID-19 on Energy Intake and Physical Activity Levels in Australian University Students. Nutrients. 2020;12(6).

12.	The Automated Self-Administered 24 Hour Dietary Assessment Tool [Internet]. National Cancer Institute. 2021. Available from: <https://epi.grants.cancer.gov/asa24/#:~:text=The%20Automated%20Self%2DAdministered%2024,also%20known%20as%20food%20diaries>.

13.	Health AIo, Welfare. The Active Australia Survey: a guide and manual for implementation, analysis and reporting. Canberra: AIHW; 2003.

14.	Tison GH, Avram R, Kuhar P, Abreau S, Marcus GM, Pletcher MJ, et al. Worldwide effect of COVID-19 on physical activity: a descriptive study. Annals of internal medicine. 2020.

15.	Luciano F, Cenacchi V, Vegro V, Pavei G. COVID-19 lockdown: physical activity, sedentary behaviour and sleep in Italian medicine students. Eur J Sport Sci. 2020:1-22.

16.	Bourdas DI, Zacharakis ED. Impact of COVID-19 Lockdown on Physical Activity in a Sample of Greek Adults. Sports (Basel). 2020;8(10).

17.	Jackson SE, Garnett C, Shahab L, Oldham M, Brown J. Association of the Covid-19 lockdown with smoking, drinking, and attempts to quit in England: an analysis of 2019-2020 data. medRxiv. 2020.

18.	Niedzwiedz CL, Green MJ, Benzeval M, Campbell D, Craig P, Demou E, et al. Mental health and health behaviours before and during the initial phase of the COVID-19 lockdown: longitudinal analyses of the UK Household Longitudinal Study. Journal of Epidemiology and Community Health. 2020:jech-2020-215060.

19.	Westrupp E, Bennett C, Berkowitz TS, Youssef G, Toumbourou J, Tucker R, et al. Child, parent, and family mental health and functioning in Australia during COVID-19: Comparison to pre-pandemic data. 2020.

20.	Sidor A, Rzymski P. Dietary Choices and Habits during COVID-19 Lockdown: Experience from Poland. Nutrients. 2020;12(6):1657.

21.	Kilian C, Rehm J, Allebeck P, Braddick F, Gual A, Barták M, et al. Alcohol consumption during the COVID-19 pandemic in Europe: a large-scale cross-sectional study in 21 countries. 2021.

22.	Yang S, Guo B, Ao L, Yang C, Zhang L, Zhou J, et al. Obesity and activity patterns before and during COVID-19 lockdown among youths in China. Clin Obes. 2020:e12416.

23.	López-Sánchez GF, López-Bueno R, Gil-Salmerón A, Zauder R, Skalska M, Jastrzębska J, et al. Comparison of physical activity levels in Spanish adults with chronic conditions before and during COVID-19 quarantine. Eur J Public Health. 2020.

24.	Katewongsa P, Widyastaria DA, Saonuam P, Haematulin N, Wongsingha N. The effects of COVID-19 pandemic on physical activity of the Thai population: Evidence from Thailand's Surveillance on Physical Activity 2020. J Sport Health Sci. 2020.

25.	Papandreou C, Arija V, Aretouli E, Tsilidis KK, Bulló M. Comparing eating behaviours, and symptoms of depression and anxiety between Spain and Greece during the COVID‐19 outbreak: Cross‐sectional analysis of two different confinement strategies. European Eating Disorders Review. 2020;28(6):836-46.

26.	Foa R, Gilbert S, Fabian MO. COVID-19 and Subjective Well-Being: Separating the Effects of Lockdowns from the Pandemic. Available at SSRN 3674080. 2020.

27.	Sibley CG, Greaves LM, Satherley N, Wilson MS, Overall NC, Lee CH, et al. Effects of the COVID-19 pandemic and nationwide lockdown on trust, attitudes toward government, and well-being. American Psychologist. 2020.

28.	Fancourt D, Steptoe A, Bu F. Trajectories of depression and anxiety during enforced isolation due to COVID-19: longitudinal analyses of 59,318 adults in the UK with and without diagnosed mental illness. medRxiv. 2020.

29.	Jacob S, Mwagiru D, Thakur I, Moghadam A, Oh T, Hsu J. Impact of societal restrictions and lockdown on trauma admissions during the COVID‐19 pandemic: a single‐centre cross‐sectional observational study. ANZ journal of surgery. 2020.

30.	Andersson C, Gerds T, Fosbøl E, Phelps M, Andersen J, Lamberts M, et al. Incidence of New-Onset and Worsening Heart Failure Before and After the COVID-19 Epidemic Lockdown in Denmark: A Nationwide Cohort Study. Circ Heart Fail. 2020;13(6):e007274.

31.	Oikonomou E, Aznaouridis K, Barbetseas J, Charalambous G, Gastouniotis I, Fotopoulos V, et al. Hospital attendance and admission trends for cardiac diseases during the COVID-19 outbreak and lockdown in Greece. Public Health. 2020;187:115-9.

32.	Ball S, Banerjee A, Berry C, Boyle JR, Bray B, Bradlow W, et al. Monitoring indirect impact of COVID-19 pandemic on services for cardiovascular diseases in the UK. Heart. 2020;106(24):1890-7.

33.	Olding J, Zisman S, Olding C, Fan K. Penetrating trauma during a global pandemic: Changing patterns in interpersonal violence, self-harm and domestic violence in the Covid-19 outbreak. The Surgeon. 2020.

34.	Qureshi AI, Huang W, Khan S, Lobanova I, Siddiq F, Gomez CR, et al. Mandated societal lockdown and road traffic accidents. Accident Analysis & Prevention. 2020;146:105747.

35.	Fahy S, Moore J, Kelly M, Flannery O, Kenny P. Analysing the variation in volume and nature of trauma presentations during COVID-19 lockdown in Ireland. Bone & joint open. 2020;1(6):261-6.

36.	Joyce L, Richardson S, McCombie A, Hamilton G, Ardagh M. Mental Health Presentations to Christchurch Hospital Emergency Department During COVID‐19 Lockdown. Emergency Medicine Australasia. 2020.

37.	Henry N, Parthiban S, Farroha A. The effect of COVID-19 lockdown on the incidence of deliberate self-harm injuries presenting to the emergency room. The International Journal of Psychiatry in Medicine. 2020:0091217420982100.

38.	Ikenberg B, Hemmer B, Dommasch M, Kanz KG, Wunderlich S, Knier B. Code Stroke Patient Referral by Emergency Medical Services During the Public COVID-19 Pandemic Lockdown. J Stroke Cerebrovasc Dis. 2020;29(11):105175.

39.	Schlachetzki F, Theek C, Hubert ND, Kilic M, Haberl RL, Linker RA, et al. Low stroke incidence in the TEMPiS telestroke network during COVID-19 pandemic - effect of lockdown on thrombolysis and thrombectomy. J Telemed Telecare. 2020:1357633x20943327.

40.	Hoyer C, Weber L, Sandikci V, Ebert A, Platten M, Szabo K. Decreased admissions and change in arrival mode in patients with cerebrovascular events during the first surge of the COVID-19 pandemic. Neurological research and practice. 2020;2(1):1-4.

41.	Frisullo G, Brunetti V, Di Iorio R, Broccolini A, Caliandro P, Monforte M, et al. Effect of lockdown on the management of ischemic stroke: an Italian experience from a COVID hospital. Neurol Sci. 2020;41(9):2309-13.

42.	Paliwal PR, Tan BYQ, Leow AST, Sibi S, Chor DWP, Chin AXY, et al. Impact of the COVID-19 pandemic on hyperacute stroke treatment: experience from a comprehensive stroke centre in Singapore. J Thromb Thrombolysis. 2020;50(3):596-603.

43.	Kristoffersen ES, Jahr SH, Faiz KW, Thommessen B, Rønning OM. Stroke admission rates before, during and after the first phase of the COVID-19 pandemic. Neurological Sciences. 2021:1-8.

44.	Kristoffersen ES, Jahr SH, Thommessen B, Rønning OM. Effect of COVID-19 pandemic on stroke admission rates in a Norwegian population. Acta Neurol Scand. 2020.

45.	Leske Stuart, Kõlves KK, Crompton D, Arensman E, de Leo D. Real-time suicide mortality data from police reports in Queensland, Australia, during the COVID-19 pandemic: an interrupted time-series analysis. Lancet Psychiatry. 2020.

46.	Lei L, Huang X, Zhang S, Yang J, Yang L, Xu M. Comparison of Prevalence and Associated Factors of Anxiety and Depression Among People Affected by versus People Unaffected by Quarantine During the COVID-19 Epidemic in Southwestern China. Med Sci Monit. 2020;26:e924609-e.

47.	González J, Moncusí-Moix A, Benitez ID, Santisteve S, Monge A, Fontiveros MA, et al. Clinical Consequences of COVID-19 Lockdown in Patients With COPD: Results of a Pre-Post Study in Spain. Chest. 2021.

48.	Mesnier J, Cottin Y, Coste P, Ferrari E, Schiele F, Lemesle G, et al. Hospital admissions for acute myocardial infarction before and after lockdown according to regional prevalence of COVID-19 and patient profile in France: a registry study. Lancet Public Health. 2020;5(10):e536-e42.

49.	Lantelme P, Couray Targe S, Metral P, Bochaton T, Ranc S, Le Bourhis Zaimi M, et al. Worrying decrease in hospital admissions for myocardial infarction during the COVID-19 pandemic. Arch Cardiovasc Dis. 2020;113(6-7):443-7.

50.	Kamine TH, Rembisz A, Barron RJ, Baldwin C, Kromer M. Decrease in Trauma Admissions with COVID-19 Pandemic. West J Emerg Med. 2020;21(4):819-22.

51.	Chen S, She R, Qin P, Kershenbaum A, Fernandez-Egea E, Nelder JR, et al. The Medium-Term Impact of COVID-19 Lockdown on Referrals to Secondary Care Mental Health Services: A Controlled Interrupted Time Series Study. Front Psychiatry. 2020;11:585915.

1. Mixing of effects, whereby the putative confounder is a cause (or proxy of a cause) of the exposure and also directly or indirectly effects the outcome independent of the exposure (here social restrictions or lock down).
2. Mismeasurement or misclassification of any of the exposure, confounders or outcome in such a way that bias is introduced to the magnitude of exposure-outcome association. In the context of COVID-19, an important source of measurement error is ‘differential measurement error of the outcome (e.g. alcohol intake) by level of the exposure’, due to people being unblinded to their exposure status (i.e. they know they are in lock-down). This is even more of a risk if the outcome measure is subjective (e.g. self-reported alcohol consumption).
3. The exposure-outcome association among the observed differs from that in the total target population of inference. For example, in COVID-19 many studies were (necessarily) rapidly constructed using snowball and internet recruitment; it is likely that people completing an online questionnaire during a lock down might have a different alcohol consumption (both current, and recalled) than someone not responding to an online questionnaire.
